# Functional Profiling of DNA Repair Pathways in Lung Cancer Patients Uncovers Radiotherapy-Induced and Cancer-Associated Alterations in Oxidative Lesion Repair

**DOI:** 10.64898/2026.01.12.26343971

**Authors:** Sneh M. Toprani, Ting Zhai, Maeve Dillon-Martin, Patrick F. Doyle, Claire Novack, David Kozono, Zachary D. Nagel

## Abstract

DNA repair capacity (DRC), particularly at the pathway level, varies among individuals. While previous studies explored DRC in relation to environmental exposures and cancer risk, few measured DRC in patient-focused cohorts and were focused on one or two repair pathways only. We comprehensively profiled DRC for all the major repair pathways and DNA lesions in 100 lung cancer patients undergoing radiotherapy (RT) using advanced Fluorescence Multiplex based Host Cell Reactivation assays in blood cells before and after RT and investigated how DRC responded to RT and was influenced by clinical variables. Variation between individuals was significant in all pathways and smaller than variation within-person. DNA glycosylase activity decreased immediately following RT and subsequently returned to baseline in patients receiving high-intensity RT during the follow-up months. Lower DRC against oxidative lesions was found in cancer patients compared to healthy controls. These results highlight oxidative DNA damage repair as a sensitive marker of RT response and cancer burden upon profiling the DNA repair landscape.

**Graphical Abstract:** 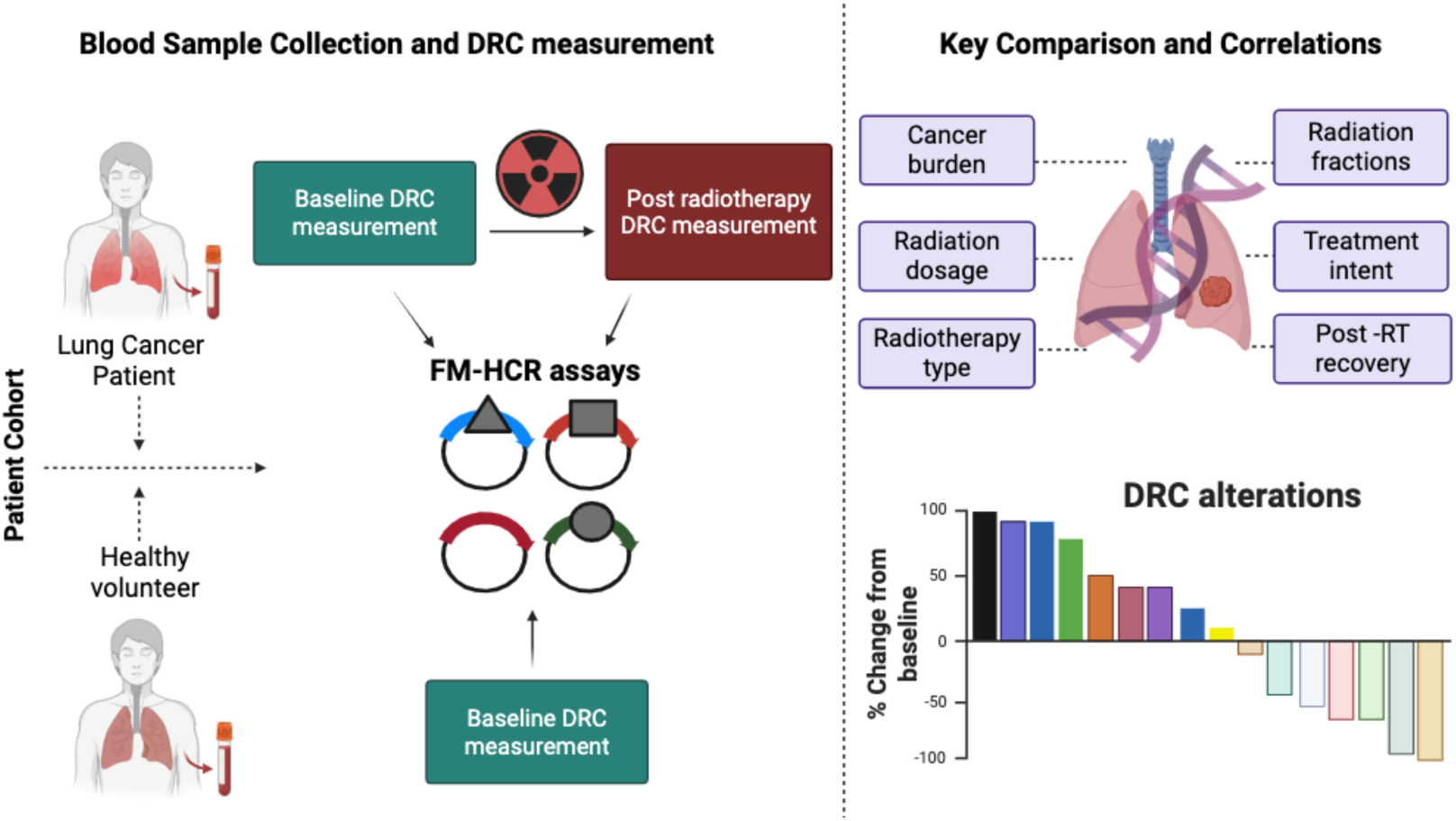

## 2. Introduction

Lung cancer remains the leading cause of cancer-related mortality, with approx. 227,000 new cases and approx. 125,000 deaths anticipated in the United States in 2025 ^1^. It arises from a complex interplay between genetics, lifestyle and environmental exposure to DNA damaging agents such as tobacco smoking, radon, air pollutants, and occupational carcinogens ^2^. DNA repair is pivotal for maintaining genomic integrity. Individuals with DNA repair capacity (DRC) deficiencies accumulate potentially carcinogenic mutations, leading to higher risk of cancer ^3^. At the same time, DRC is inversely associated with cancer patient survival following treatment with DNA damaging chemotherapy, potentially indicating that higher DRC in lymphocytes is a marker for higher DRC in tumors ^4^. Although multiple studies have linked lower DRC to higher lung cancer susceptibility ^5–8^, the underlying causes of inter-individual differences in DRC and the extent to which they are influenced by treatment and cancer burden remained incompletely understood. Elucidating both baseline and longitudinal variation in DRC ^9–13^ is therefore expected to offer critical insights into lung cancer susceptibility and prognosis.

In addition to cancer risk, DRC is expected to play an important role in determining individual’s susceptibility to the side effects of radiotherapy (RT). Lung cancer patients exhibit significantly reduced DRC compared to healthy individuals, particularly in the nucleotide excision repair (NER) ^8,14–17^ and base excision repair (BER) ^5,18^ pathways. However, other critical DNA damages and repair pathways have been less studied due to assay limitations that precluded simultaneous assessment of multiple DNA repair pathways. RT achieves its therapeutic effects largely by inducing diverse types of DNA base damage and strand breaks in tumor cells. These diverse DNA lesions are repaired by multiple repair pathways. RT also damages DNA in non-cancerous cells, leading to normal tissue toxicity ^19^. Toxicity might be predicted from an individual’s DRC, but it is not yet clear whether DRC remains stable over time in patients or is modified by patient demographics, clinical factors or RT parameters. Literature lacks longitudinal mapping of DRC across major DNA repair pathways which is needed to determine the translational potential of DRC assays for informing strategies to improve patient outcomes ^20^.

Blood-based assays provide a minimally invasive method to assess tumor activity and treatment responses in lung cancer patients ^4,5,8,14,15,18,21–24^. Functional assays hold particular promise for measuring human variation ^25^. Few individuals have germline mutations in DNA repair genes that are associated with altered function^26^, and so DNA sequencing as an approach is not expected to capture this variation in most cases, nor would it capture epigenetic or environmental changes. Strong correlations between 8-oxoguanine DNA glycosylase (OGG) activity in blood cells and lung cells from the same patients reinforce the validity of blood cells as a surrogate for other tissues ^18^. Nonetheless, most genome integrity assays (e.g., mutagen sensitivity, immunofluorescence) are restricted to specific repair enzymes or pathways ^27,28^, and have limitations in assessing broader DNA repair networks ^29,30^. Previous DRC studies in lung cancer patients only focused on one or two DNA repair enzymes and host cell reactivation assays were only used in mitogen stimulated peripheral blood mononuclear cells (PBMCs) rather than quiescent live cells, thus introducing a potential source of sample variability ^5,14,15,24,31–35^. To address these gaps, we utilized advanced high-throughput Fluorescence Multiplex based Host Cell Reactivation (FM-HCR) assays, optimized for simultaneously measuring DRC across all major DNA repair pathways ^13,36,37^. This method has been extensively validated in our previous work on cell lines ^36,38–41^ and PBMCs ^10,13,25,30,37^, offering a robust and reproducible platform for evaluating the DNA repair landscape in lung cancer patients. We measured DRC in PBMC collected before and after RT and integrated these functional data with patient clinical information. We also obtained PBMCs from a demographically matched group of cancer-free individuals. Our results indicate that DNA repair landscapes are influenced by treatment but are generally stable and significantly different between individuals. DRC in some repair pathways is also suppressed in cancer cases relative to controls. Such novel comprehensive studies about all major DNA repair pathways at individual level could be transformative for identifying and tailoring treatment and radiation induced side effects for patients whose DRC put them at higher risk for RT related adverse health outcomes. Taken together, our findings underscore the potential for personalized cancer treatment and prevention strategies tailored to individuals based on their DRC profiles.

## 3. Methods

### 3.1. Study Design and Patient Cohort

This study enrolled adult lung cancer patients at the Radiation Oncology Department of Brigham and Women’s Hospital between 2019 and 2022 in Boston, Massachusetts, USA. Eligible participants included those with primary or metastatic tumors receiving RT targeting sites within the lung (Figure 1). Patients were excluded if they were deemed temporarily or permanently unfit to consent to research by their treating radiation oncologist or if they were medically unstable (i.e. ICU admitted) at the time of RT mapping. Additionally, patients with insufficient blood cell counts were excluded, as this precluded adequate blood sampling for downstream analyses. A total of 100 patients were included in the final cohort. All participants provided written informed consent prior to enrollment. The study was approved by the Institutional Review Board at Brigham Women’s Hospital (MGB IRB #2016P001582).

**Figure 1.**
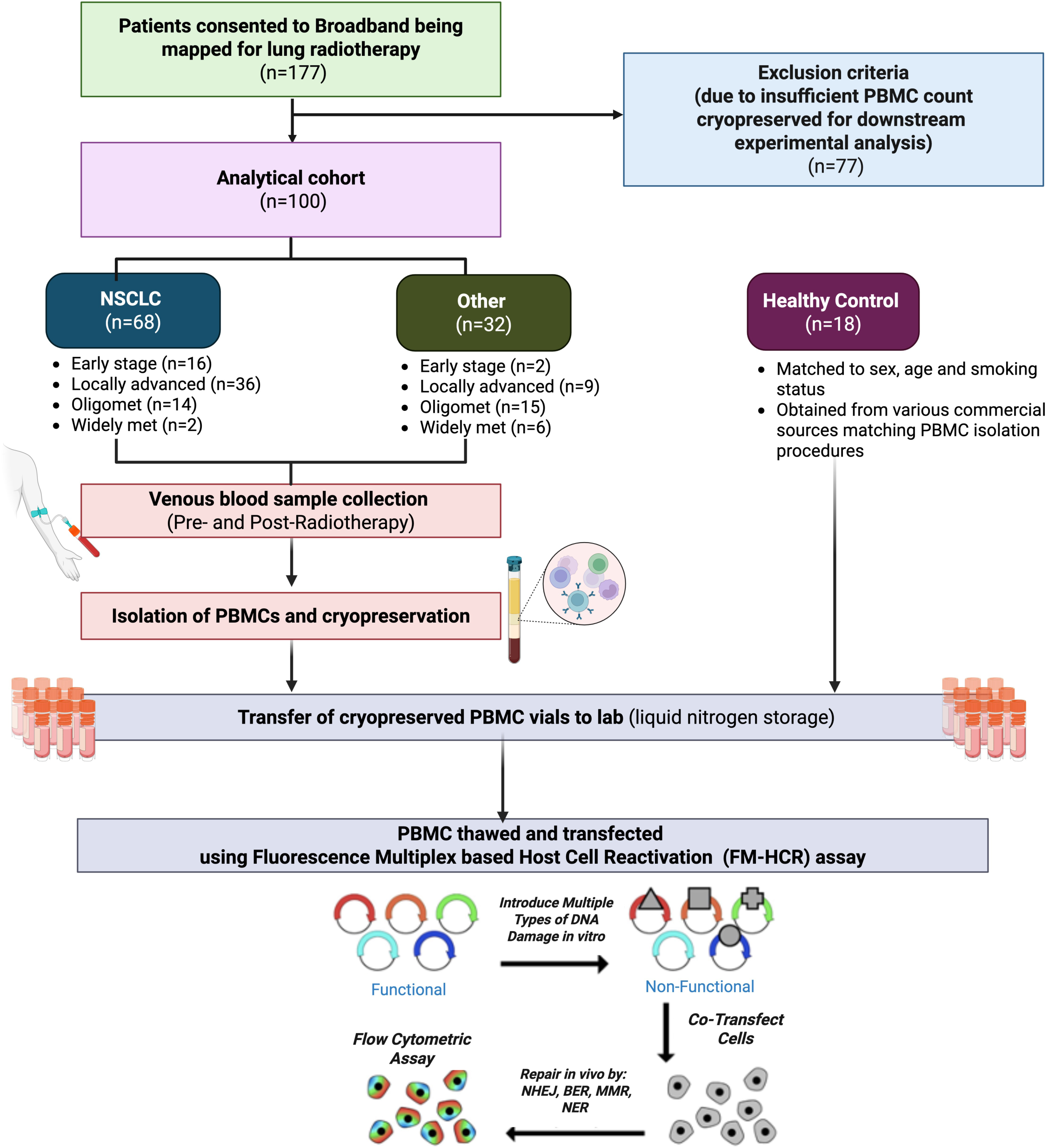
Schematic of the study design. Patients receiving thoracic radiotherapy (RT) for lung tumors were enrolled in the Broadband Study after providing informed consent. Venous blood samples were collected from each patient at two time-points: pre-RT and post-RT. Peripheral blood mononuclear cells (PBMCs) were isolated using density gradient centrifugation, cryopreserved, and subsequently transferred to the Harvard laboratory and stored in liquid nitrogen. From 177 patients with cryopreserved PBMCs, 100 patient samples were selected based on PBMC cell count and experimental requirements, blinded and randomly distributed into 26 groups. PBMCs were thawed and transfected to assess DNA repair capacity using Fluorescence Multiplex based Host Cell Reactivation assays. Additionally, PBMC samples from healthy controls matched based on patient demographics (age, sex and smoking status) were obtained from commercial sources. Control PBMCs from commercial sources underwent identical isolation and cryopreservation procedures.

Blood samples were sequentially collected in EDTA coated vacutainers (BD K_2_EDTA vacutainers, cat no. 367863) from patients before (baseline) and post RT (post-RT). The follow-up period ranged from within one month to over a year post-RT. For this study, we focused on two key visits: one before RT, and the earliest post-RT visit. Samples were processed and characterized to incorporate biological data into analyses of RT response.

For case-control matched studies, a subset of the patient cohort (post-RT timepoint) was matched with healthy individuals based on age and sex. Peripheral blood mononuclear cells (PBMCs) from these control (healthy) individuals were obtained from commercial sources (SMEBio, USA; Cellular Technology Limited, Ohio; Precision for Medicine, Massachusetts), with the same PBMC isolation and cryopreservation protocol applied.

### 3.2. Clinical Information Collection

Comprehensive RT information, including treatment dates, technique, prescribed and delivered doses and fractions, target site, and prior RT history, was obtained from on-site RT records. Patient demographics, such as age, sex, race, and smoking history, were collected via standardized questionnaires at enrollment. Primary cancer diagnosis, histology, and staging information were obtained from cancer registry and medical records. Treatment history including previous or concurrent cancer therapies such as chemotherapy, targeted therapy, and immunotherapy were extracted from medical records. The clinical data underwent quality control conducted by a team of radiation oncologists and trained staff.

Patients were categorized into two groups based on tumor histology: non-small cell lung cancer (NSCLC) and other cancers, the latter comprising small cell lung cancer (SCLC) and secondary (metastatic) cancers within the lung. Cancer stage was classified into four categories - early stage, locally advanced, oligometastatic, and widely metastatic - based on AJCC 8th Edition staging criteria and treatment intent. RT type was further grouped into stereotactic body RT (SBRT), radical (curative) standard fractionated therapy, and palliative therapy, classified by treatment technique, total dose, and fractionation.

### 3.3. Sample collection

Blood samples collected from the patients were sent to Clinical Research Facility at Dana Farber Cancer Institute, Boston, Massachusetts, USA. Peripheral blood mononuclear cells (PBMCs) were isolated using density gradient centrifugation with Lymphoprep (StemCell Technologies, Cat no. 07811) in SepMate tubes (StemCell Technologies, Cat no. 85460), following the manufacturer’s protocol ^37,80^. Cell viability and quantity were assessed using the Trypan blue dye exclusion assay using Vi-Cell XR Cell counter, Beckman Coulter (Cat no. TOC Analyzer QbD1200). After cell counting, PBMCs were washed with 1X phosphate buffer saline (PBS; Gibco, Cat no. 14190144) containing 2% fetal bovine serum (FBS; Gibco, Cat no. 10437028) at 400g for 8 mins. The washed cells were then resuspended in 1 ml of ice-cold RPMI 1640 medium (Gibco, Cat no. 11875085) supplemented with 20% FBS and 10% dimethyl sulfoxide (DMSO, VWR, Cat no. IC0219605590). Cell suspensions were aliquoted into 1.8 ml cryovials (ThermoFisher Scientific, Cat no. 347627) (with deidentified barcodes) with each vial containing up to 10 million cells/ml. For controlled freezing, the cryovials were placed in a Mr. Frosty freezing container (Nalgene ThermoFisher Scientific, Cat no. NL51000001) which affords a cooling rate of approximately −1°C/min for 24 hr. Thereafter, the vials were transferred to cryoboxes and stored at −80°C freezer in Biospecimen Repository Core, Dana Farber Cancer Institute. To minimize batch effects, all samples were processed simultaneously, and the tubes were transferred (three month prior to the start of the experiment) to an automated controlled liquid nitrogen Dewar at Harvard T.H Chan School of Public Health, Boston, Massachusetts, USA.

### 3.4. Cryopreserved PBMC processing for downstream analysis

Cryopreserved PBMC samples were de-identified and randomly divided into a total of 26 batches, with each batch consisted of samples collected at two timepoints (before and post-therapy) from ∼8 patients being processed on the same day, whereas for case-control matched samples, each batch consisted of 8 samples of age and sex matched case-control pairs. Each PBMC vial was quickly thawed in a 37°C water bath, then resuspended in ice cold RPMI 1640 medium supplemented with 20% FBS. The thawed PBMCs were washed twice with 1X PBS containing 2% FBS at 400 g for 8 mins to remove any residual cryoprotectant. Following cell counting, PBMCs were suspended at density of 1 million cells/ml in pre-warmed RPMI medium containing 20% FBS for 6 hr at 37°C with 5% CO_2_.

### 3.5. Assessing DRC using Fluorescence Multiplex based Host Reactivation (FM-HCR) assay

#### Plasmid reporter and cocktail preparation

DRC was assessed using FM-HCR assays. The detailed methodology for this assay and the preparation of plasmid-based lesion has been reported previously, ^10,36,37,81^ but is summarized here. Cocktails of fluorescent reporter plasmids with defined DNA lesions were assembled as outlined in Table 1, which provides detailed information on the DNA lesions incorporated into the reporter plasmids, repair pathways measured, and fluorochromes used. Each cocktail included a transfection control plasmid (pMax_mPlum for the cocktail “damage 1”, pMax_GFP for “damage 2”; pMax_mOrange for “damage 3”) to normalize transfection efficiency (TE). A carrier plasmid that does not express a fluorescent protein (pMax_mCherry_Δ-Δ-CMV) was also included in each cocktail to enhance TE. FM-HCR plasmids reported the ability of cells to repair several DNA lesions, including uracil (**U:G**) lesions that are repaired by uracil DNA glycosylases (UDG, TDG, SMUG1, and MBD4); hypoxanthine (**Hx:T**) lesions repaired by methylpurine DNA glycosylase (MPG); **8oxoG:C** lesions repaired by various DNA glycosylases (OGG1, NEIL1, and NEIL2), and **A:8oxoG** lesions repaired by MUTYH DNA glycosylase. For these four reporters, DRC is inversely related to the fluorescent reporter protein expression because signal arises from transcriptional mutagenesis of unrepaired DNA lesions. For the remaining reporter plasmids, DRC is directly proportional to fluorescent protein expression. These included double strand breaks repaired by non-homologous end joining **(NHEJ)**, tetrahydrofuran (abasic site analog, THF) lesions repaired by long patch-base excision repair (**LP-BER**); single base mismatches (G:G) repaired by mismatch repair **(MMR)**; ultraviolet light-induced damage repaired by nucleotide excision repair **(NER)**, and ionizing radiation (**IR**)-induced lesions that are repaired by multiple DNA repair pathways.

**Table 1:** Composition of plasmid cocktails used in this study: Four plasmid cocktails (undamaged 1-3, damaged 1, damaged 2, and damage 3) of four colors (BFP, GFP, mOrange, and mPlum) were prepared to measure the repair capacity of cells for nine DNA repair lesions/pathways. pMax backbone plasmid was used for this study. Details of the plasmid fluorochrome reporter, filter, and flow channel was used to gate the fluorescent reporter events. The amount (in nanograms) of each plasmid reporting for a particular pathway of interest is mentioned. pMax_mPlum (in damaged cocktail 1), pMax_GFP (in damaged cocktail 2), and pMax_mOrange (in damaged cocktail 3) were used as transfection controls.

| Fluorochrome of reporter plasmid (filter, flow channel) | Undamaged cocktail (1-3) |  |  | Damaged cocktail (1) |  |  | Damaged cocktail (2) |  |  | Damaged cocktail (3) |  |  |
| --- | --- | --- | --- | --- | --- | --- | --- | --- | --- | --- | --- | --- |
|  | *Plasmid* | *Qty (ng)* | *Measure* | *Plasmid* | *Qty (ng)* | *Measure* | *Plasmid* | *Qty (ng)* | *Measure* | *Plasmid* | *Qty (ng)* | *Measure* |
| BFP<br>(440/50, VL-1) | pMax_BFP | 30 | ---- | BFP_NHEJ | 50 | NHEJ | BFP_U | 30 | Uracil lesion | pMax_BFP_IR (3000Gy) | 100 | Ionizing radiation (IR) induced damage |
| GFP<br>(530/30, BL-1) | pMax_GFP | 10 | ----- | GFP_THF | 10 | Long patch BER (LP-BER) | pMax_GFP | 10 | Control | GFP_Hx:T | 20 | Hypo-xanthine (Hx:T) lesion |
| mOrange<br>(585/16, YL-1) | pMax_mOrange | 20 | ----- | mOrange_GG | 20 | MMR | mOrange_8oxoG-C | 60 | 8oxoG:C lesion | pMax_mOrange | 20 | Control |
| mPlum<br>(695/40, YL-3) | pMax_mPlum | 50 | ----- | pMax_mPlum | 50 | Control | mPlum_A-8oxoG | 200 | A:8oxoG lesion | mPlum_UV 800J/m2 | 100 | NER |
| Carrier plasmid DNA (non-fluorescent) | pMax-mCherry-ΔΔ-CMV | 1000 | ----- | pMax-mCherry-ΔΔ-CMV | 1000 | ----- | pMax-mCherry-ΔΔ-CMV | 1000 | ----- | pMax-mCherry-ΔΔ-CMV | 1000 | ----- |

#### Transfection of PBMCs with plasmid cocktails

After a 6-hr incubation, PBMCs were collected by centrifugation at 400 g for 8 mins and washed with 1XPBS containing 2% FBS. For each transfection, 2X10^5 cells were suspended in a 10 μL volume of T buffer provided by the manufacturer and transfected using a *Neon transfection kit* (Thermofisher Scientific, Cat no. MPK1096). Five aliquots of 10 μL each were distributed into individual tubes of a 0.2 mL (8-tube) PCR strip: four aliquots were used for transfection with different plasmid cocktail (containing either damage or undamaged plasmids), and one aliquot served as the mock un-transfected control. Cells were combined with a 1.5μL volume of plasmid cocktail composed in TE buffer as detailed in Table 2. Electroporation was performed using the Neon Transfection System (Thermofisher Scientific, Cat no. MPK5000) at 2250V with a single 20ms pulse. Single color control transfections with 200 ng of damage-free reporter plasmids were performed to establish gating, determine voltage settings, and set compensation (**Fig S1-S2**). The transfected cells were transferred to pre-warmed RPMI medium supplemented with 20% FBS in a 24-well tissue culture plate and cultured at 37°C with 5% CO_2_ in a humidified atmosphere for 18 hr.

**Table 2.** Baseline characteristics of enrolled patients.

| Variable | Summary |
| --- | --- |
| Age (years) | 66 (61-73) |
| Female | 49 |
| White race | 90 |
| Smoking |  |
| Never | 21 |
| Former | 69 |
| Current | 9 |
| NSCLC | 68 |
| Stage |  |
| Early stage | 18 |
| Locally advanced | 45 |
| Oligomet | 29 |
| Widely met | 8 |
| Therapy intent |  |
| SBRT | 24 |
| Radical | 71 |
| Palliative | 5 |
| Dosage (Gy) | 60 (54-60) |
| Fractions | 27 (12-30) |
| Prior radiotherapy |  |
| Any | 15 |
| Within 6 months | 6 |
| Prior chemotherapy | 31 |
| Prior immunotherapy | 13 |
| Prior targeted therapy | 17 |

Continuous variables are summarized with median (interquartile range), categorical variables are summarized with counts.
Abbreviations: NSCLC = non-small cell lung cancer, SBRT = stereotactic body radiotherapy

#### Flow cytometry and analysis

Following the 18 hr post-transfection incubation period, cells were thoroughly resuspended by trituration and transferred to flow cytometry tubes (VWR, Cat no. 60818-496) for analysis using an Attune NxT Flow Cytometer (Thermofisher Scientific, Cat no. A29001). Cell debris and doublets were excluded based on their side and forward scatter characteristics. Data were processed using Attune NxT software and Microsoft Excel, in accordance with a detailed analysis protocol^36^. Briefly, to ensures quality-control, a minimum of 30 fluorescent positive events was established for data inclusion in the FM-HCR analysis. The fluorescent cell count was subsequently multiplied by the mean fluorescence intensity for both damaged and undamaged cocktails. For each plasmid cocktail used in the transfection, the TE of each reporter plasmid was normalized to the transfection control to account for variation in TE. Overall, repair capacity was quantified as the percent ratio of reporter expression from damaged reporter plasmid to the corresponding undamaged reporter plasmid^36–38^.

### 3.6. Statistical analysis

Detailed data pre-processing steps, including batch effect correction and data standardization, have been described in detail in previous work ^13^. Briefly, raw reporter expression data were log-transformed and scaled to minimize the impact from outliers and approximate a normal distribution for subsequent analysis. For the U:G, Hx:T, A:8oxoG and 8oxoG:C repair data, the signs of the standardized values were inverted so that higher values correspond to higher repair capacity. The data presented in the results were expressed as z-scores with standard deviations as units to reflect differences between individuals at the cohort level. Batch effect correction was performed using the *ComBat* algorithm ^82^, adjusting for covariates including age, sex, smoking status, sample collection time point, and transfection efficiency, to preserve biological variation while minimizing technical variability across batches.

Generalized linear regression models were employed when analyzing associations between patient characteristics and baseline DRC. A comprehensive list of variables was explored through univariate regression, including age, sex, race (Black, Asian, vs White), smoking status (current, former vs never smoker), primary cancer diagnosis (NSCLC vs non-NSCLC), cancer stage (widely metastatic, oligo-metastatic, locally advanced vs early stage), number of primary diagnoses, RT type (palliative, radical vs SBRT), prior RT, prior chemotherapy, prior immunotherapy, prior targeted therapy. These variables were selected based on prior evidence or biological hypothesis regarding their potential role in influencing DRC. Trend tests were performed by treating cancer stage as a continuous variable to evaluate association with increased stage severity.

To evaluate the response of DRC to RT, linear mixed-effects models were used to account for within-individual correlations due to repeated measures. These models assessed the main effect of therapy status (post-RT vs pre-RT), while controlling for relevant covariates including age, sex, smoking status, cancer stage and type, RT type, dosage, and fraction, and other therapies received prior to or during sample collection. For analyzing changes in DRC over post-RT follow-up months, we applied generalized linear regression models with a Gamma distribution to accommodate the right-skewed distribution of DRC changes measured at various months post-RT and bounded at t=0. *Pearson’s* correlation tests were performed to evaluate the relationships among different DRC pathways measured simultaneously, with multiple testing corrections applied using the Holm method.

Statistical analyses were performed in *R* version 4.3.3. All p-values were two-sided, with a cut-off of <0.05 for statistical significance.

## 4. Results

To understand whether cancer burden or radiotherapy (RT) affects individual’s DNA repair capacity (DRC), DRC was comprehensively profiled using advanced high-throughput Fluorescence Multiplex based Host Cell Reactivation (FM-HCR) assays in unstimulated PBMCs obtained from lung cancer patients at baseline (pre-RT) and post-RT [Figure 1-2A]. DRC was further measured in matched cancer-free controls (healthy volunteers). FM-HCR is a well-established functional based approach that simultaneously measures DRC in all major DNA repair pathways including non-homologous end joining (NHEJ), nucleotide excision repair (NER), direct reversal (DR), mismatch repair (MMR), homologous recombination (HR) and base excision repair (BER) ^36,37^. It’s reproducibility and ability to robustly detect small interindividual differences in DRC at large population level have been validated in both quiescent and proliferating PBMCs ^13,30,37^. Reporter plasmids, each containing specific DNA lesions, were transiently transfected into quiescent PBMCs to evaluate repair capacities across multiple DNA repair pathways as detailed in **Table 1., Figure S1-S2, and the methodology section**.

**Figure 2.**
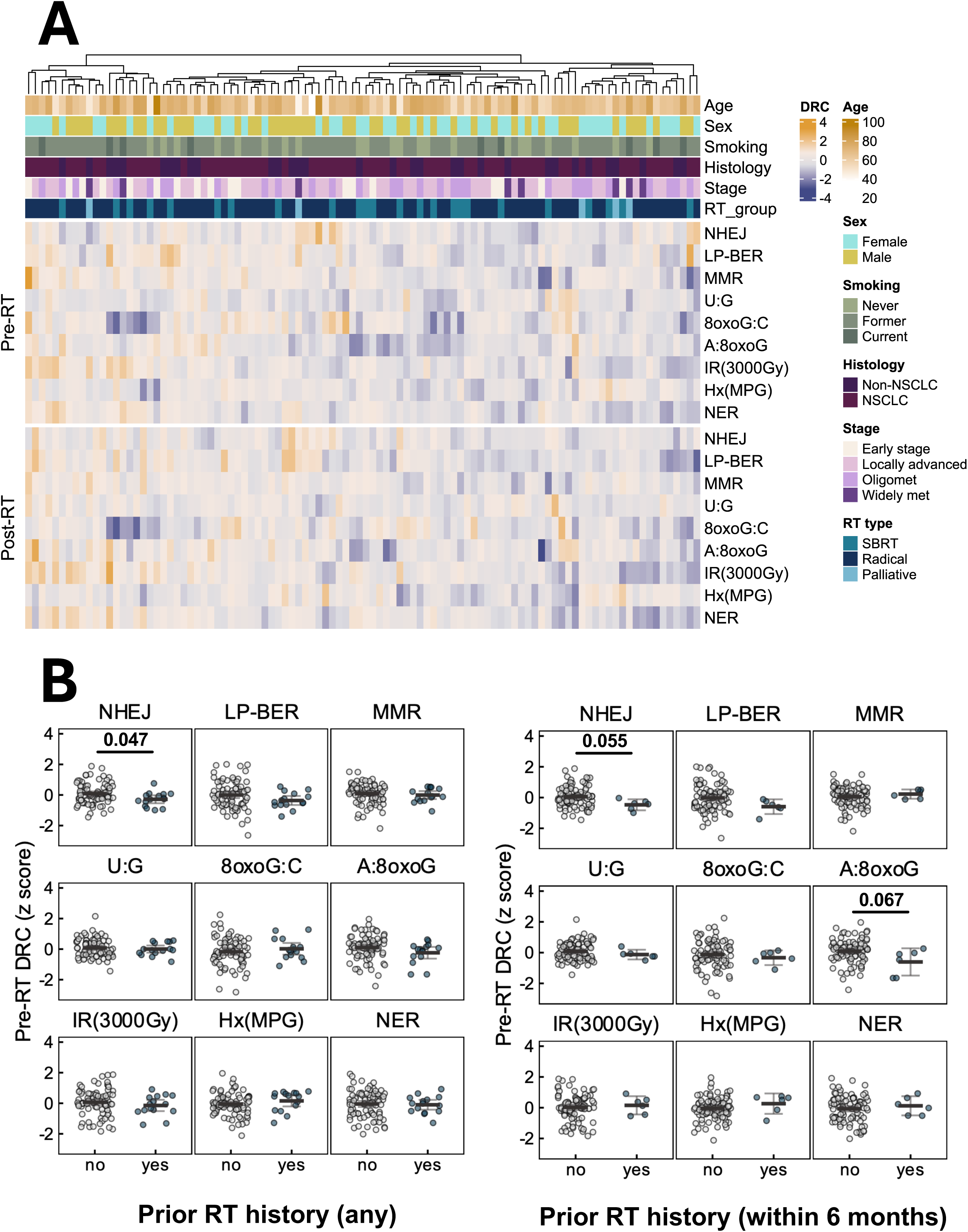
DNA repair capacity landscape in lung cancer patients before and after radiotherapy. **(A)** Landscape of DNA repair capacity (DRC) in patients across multiple DNA repair pathways, including NHEJ, repair of a THF abasic site analog (LP-BER), MMR, NER, IR-induced damage, and DNA glycosylase-dependent initiation of BER for a U:G lesion, Hx:T lesion, 8oxoG:C lesion, or A:8oxoG lesion. DRC was measured before radiotherapy (pre-RT) and after radiotherapy (post-RT). DRC values were z-scored and row-scaled for visualization. Columns represent patients; rows represent pathways. Annotations include patient age, sex (male, female), smoking status (never, former, current), cancer type (non-small cell lung cancer [NSCLC], non-NSCLC), cancer stage (early stage, locally advanced, oligometastatic, and widely metastatic), and RT type (stereotactic body radiation therapy [SBRT], radical, and palliative radiotherapy). **(B)** Influence of prior RT on pre-RT DRC. Baseline DRC z-scores (y-axis) are compared between patients with and without prior RT history (left panel), and between those with and without RT exposure within the previous 6 months (right panel) preceding the date of the baseline sample collection visit. Group comparisons with statistical significance or borderline significance from generalized linear models are marked with corresponding p-values.

### 4.1. Patient characteristics

A cohort of 100 cancer patients treated with thoracic RT was enrolled [Figure 1], with a median age of 66 years (interquartile range [IQR]: 61 – 73 years). Among these, 49 were female [Table 2]. The majority of participants were White (n = 90), and a substantial proportion were former smokers (n = 69). Of the cohort, 68 patients had a primary diagnosis of NSCLC; other cancer types mainly included SCLC, mesothelioma, colorectal cancer, and endometrial cancer [Figure S3A]. 18 patients were diagnosed with early-stage disease and 45 patients were diagnosed with locally advanced disease. 71 patients were treated with radical RT. The median RT dose delivered was 60 Gy (IQR: 54 – 60 Gy), administered over a median of 27 fractions (IQR: 12 – 30), most commonly either 3–5 fractions for stereotactic body RT (SBRT) or 30 fractions for radical standard fractionated RT. The median follow-up period since RT completion was 2.8 months (IQR: 1.3 – 5.4 months). Notably, 15 patients had received RT prior to enrollment from a previous treatment course, with 6 among them receiving RT within 6 months preceding the baseline assessment.

### 4.2. Cancer stage shapes baseline DRC of an individual

At baseline, generalized linear models were used to discover patient characteristics associated with DRC. Patients who had received RT within the past 6 months exhibited a borderline significant trend towards lower **A:8oxoG** lesion repair capacity compared to those without recent RT (estimate = −0.664 standard deviation [s.d.], p = 0.067), but the same trend was not observed in those with prior RT beyond 6 months [Figure 2B]. Additionally, baseline samples collected from patients with prior RT showed reduced **NHEJ** repair capacity (estimate = −0.367 s.d., p = 0.047). Conversely, prior immunotherapy (estimate = 0.483 s.d., p = 0.022) and chemotherapy (estimate = 0.312 s.d., p = 0.044) were associated with enhanced repair of **Hx:T** lesions.

Further analysis revealed that baseline DRC of the patients did not significantly differ across cancer type (NSCLC vs non-NSCLC) or RT type (SBRT, radical standard fractionated RT, and palliative RT). The non-NSCLC group was small and heterogenous; thus, no formal statistical tests were performed among the subcategories [Figure S3B]. However, significant variation in baseline DRC was observed across cancer stages [Figure 3A-C]. **Ionizing radiation (IR) induced damage** repair capacity was significantly higher in early-stage patients compared to locally advanced (estimate = −0.490 s.d., p = 0.037) and widely metastatic (estimate = −0.796 s.d., p = 0.024) patients [Table S1]. Furthermore, a borderline significant progressive decline in **IR induced damage** repair capacity was observed with increasing metastatic burden (trend test p = 0.053, per one stage increment from early to widely metastatic stage). In contrast, repair activity was significantly higher in patients at locally advanced stage compared to patients in early stage (estimate = 0.441 s.d., p = 0.017) and oligometastatic stages (estimate = 0.405 s.d., p = 0.010). No significant associations were found between repair capacity and patient demographics, including age, sex, and smoking history [Table S1, Figure S4].

**Figure 3.**
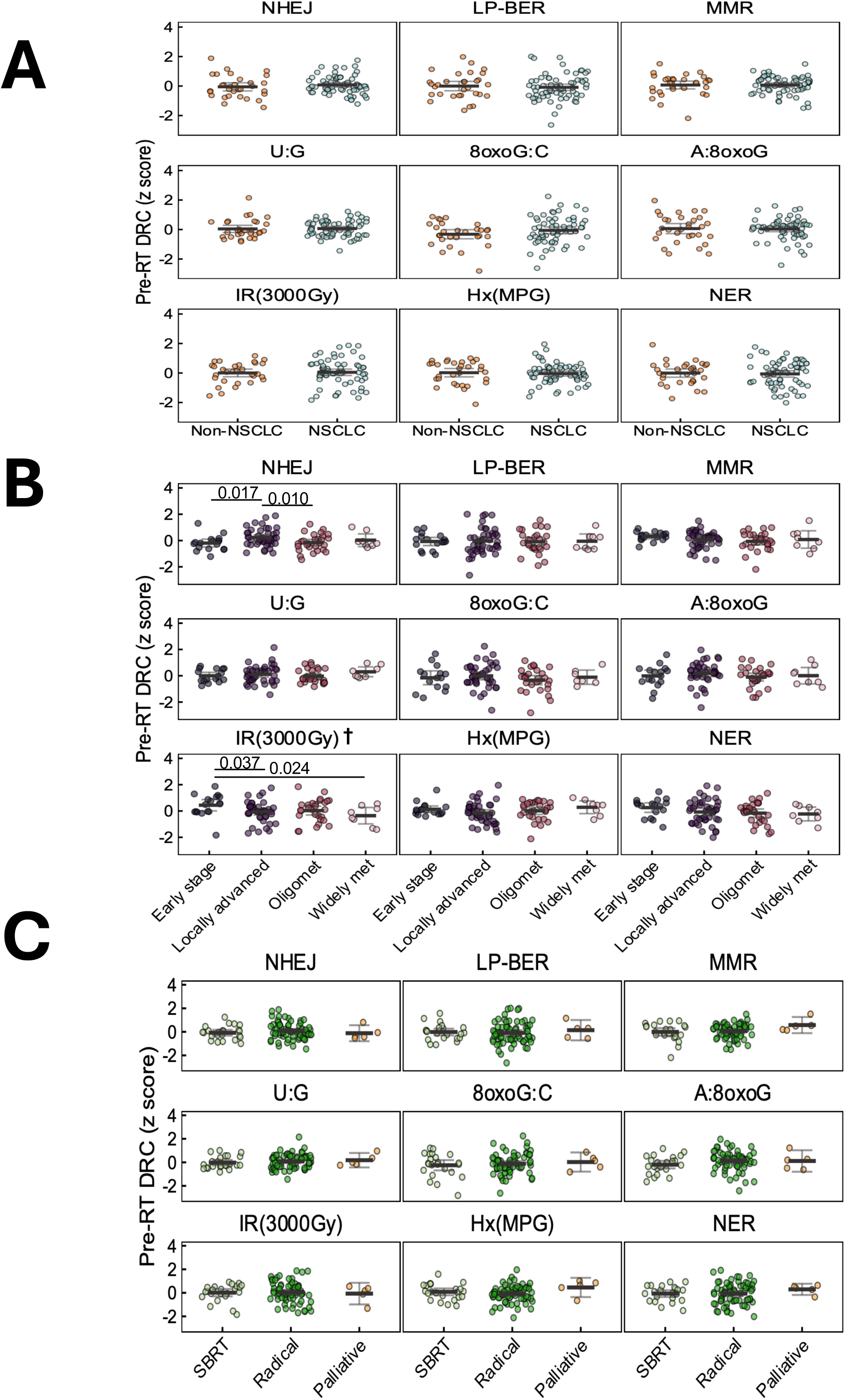
Variation in baseline DNA repair capacity by clinical characteristics. Plots report Z-scored pre-RT DNA repair capacity (DRC) across multiple pathways. DRC distributions are shown by **(A)** cancer type (non-small cell lung cancer [NSCLC] and non-NSCLC), **(B)** cancer stage (early stage, locally advanced, oligometastatic, and widely metastatic), and **(C)** RT type and treatment intent (stereotactic body radiation therapy [SBRT], radical, and palliative radiotherapy). Statistically significant group differences from generalized linear models are annotated with p-values. †p<0.05 for trend tests treating cancer stage as a continuous variable to assess dose-response relationships with stage severity.

### 4.3. Pathway-specific changes in DRC post-RT

Linear mixed effect models revealed significant RT-associated decreases in patients’ capacity to repair **A:8oxoG** lesions (estimate = −0.175 s.d., p = 0.038) and **uracil (U:G)** lesion (estimate = −0.170 s.d., p = 0.025). These decreases remained significant after adjusting for cancer stage, RT type, patient demographics, and other cancer treatments administered prior to or during RT [Figure 4A, Table S2].

**Figure 4.**
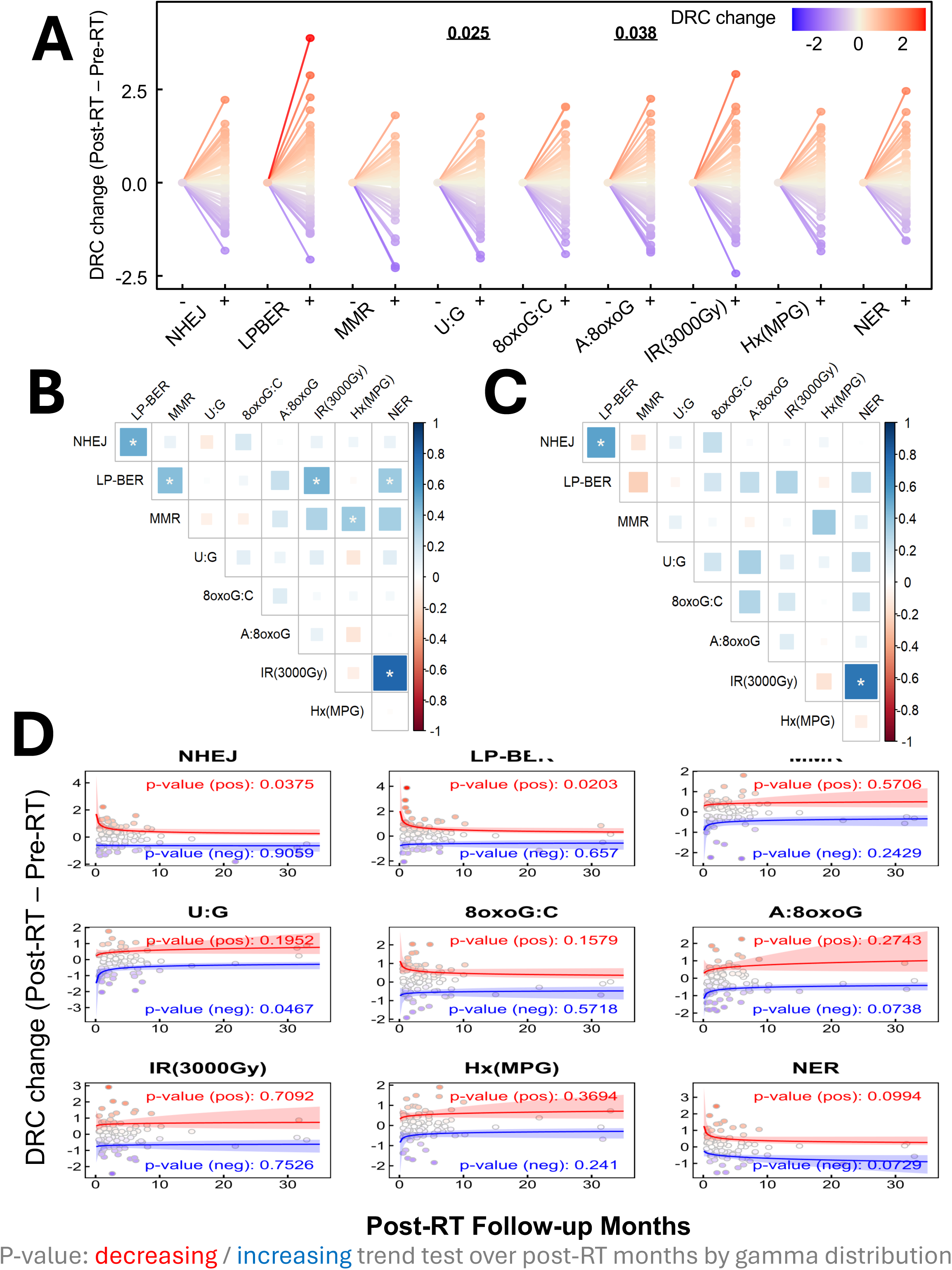
Distinct radiotherapy-induced changes in DNA repair capacity. **(A)** Patient-specific responses to radiotherapy (RT) were quantified by calculating the differences in DNA repair capacity (DRC) between post-RT (+) and pre-RT (-) timepoints across multiple pathways. Statistical significance of RT-induced changes was assessed using linear mixed-effect models adjusted for age, sex, smoking status, cancer stage, cancer type, RT type, and other therapies received prior to or during sample collection. The color gradient ranges from blue (indicating lower DRC changes) to red (indicating higher DRC changes). **(B)** Correlation matrix showing inter-pathway relationships between DRC values in pre-RT samples. **(C)** Correlation matrix showing inter-pathway relationships between DRC changes (post-RT minus pre-RT). Pearsons correlation coefficients are shown; *p<0.01 after multiple-testing adjustment. **(D)** Time-trend analysis of DRC changes (post-RT minus pre-RT) plotted against months elapsed between completion of radiotherapy and follow-up. Separate trend tests were performed for increasing (positive [pos] in red) and decreasing (negative [neg] in blue) trajectories using generalized linear models with a Gamma distribution. P-values represent the result of a trend test for decreasing or increasing DRC over post-RT months.

There were strong correlations observed between **IR induced damage** and **NER** capacities both before RT (Pearson’s R = 0.79, p <0.001) [Figure 4B] and in response to RT (Pearson’s R = 0.73, p <0.001) [Figure 4C], indicating potential co-regulation of the pathways responsible for repair of IR-induced DNA damage and UV-induced DNA damage. Moderate correlations were also evident at baseline between MMR and NER, as well as between long patch-BER (LP-BER) and NHEJ, MMR, IR induced damage, and NER [Table S3], suggesting some amount of DNA repair regulation in cancer patients.

Furthermore, significant return to baseline trends were found when we fit curves to DRC and post-RT sample collection time data for patients whose **NHEJ** (p = 0.038) and **LP-BER** (p = 0.020) repair capacities were higher post-RT, as well as in those whose **U:G** lesion (p = 0.047) repair capacity was lower post-RT [Figure 4D]. Although these trends are based on different individuals whose DRC was sampled at various times post-RT, the associations imply that changes in DRC at the individual level are largest immediately post therapy and then decay towards baseline DRC.

### 4.4. Impact of total dose and dose fractionation on RT-induced DRC changes

We further evaluated how RT treatment parameters influenced the observed RT effect on DRC by stratifying patients based on RT dosage and fraction schedule. Patients who received higher dosages (> 60Gy) showed a significant RT-induced decrease in **A:8oxoG** lesion repair capacity (estimate = −0.518 s.d., p = 0.008) [Figure 5A], even after adjusting for the number of fractions. Similarly, patients who received a higher number of RT fractions (> 20 fractions) exhibited significant RT-induced decline in **A:8oxoG** lesion repair capacity (estimate = −0.334 s.d., p = 0.005) [Figure 5B], after adjusting for RT dosage.

**Figure 5:**
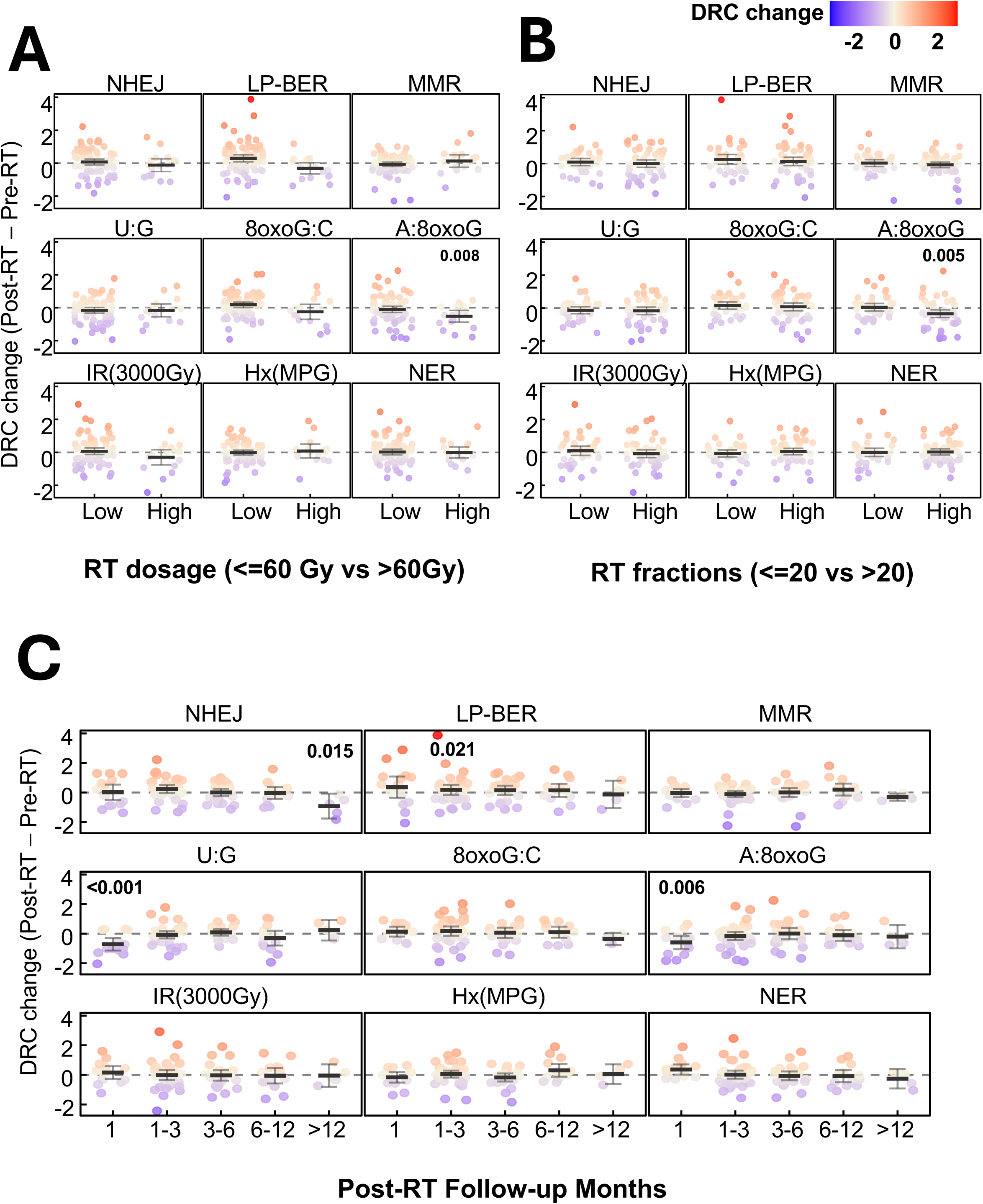
Influence of radiotherapy dosage, fractionation, and time since completion of radiotherapy on radiotherapy-induced changes in DNA repair capacity (DRC). Radiotherapy (RT)-induced changes in DNA repair capacity (DRC), calculated as post-RT minus pre-RT values, were evaluated across multiple DNA repair pathways. **(A)** Patients were stratified by radiotherapy dosage into low (<=60Gy) and high (>60Gy) dosage groups. **(B)** Patients were stratified by the number of radiation fractions received: low (<=20) vs high (>20). For both panels, p-values indicate the statistical significance of RT effects on DRC, as determined by linear mixed-effects models adjusted for the other variable (dosage or fraction) to evaluate independent associations. **(C)** DRC changes were assessed by month of post-RT follow-up to evaluate temporal patterns of repair response. P-values reflect the significance of RT effects from mixed-effects models. For all panels, each subgroup is presented with mean values and 95% confidence intervals (error bars). The color gradient ranges from blue (indicating lower DRC changes) to red (indicating higher DRC changes).

Temporal analysis of post-RT visits revealed that the majority of DRC alterations occurred shortly after treatment [Figure 5C; Table S4]. Notably, both **U:G** lesion (estimate = - 0.527 s.d., p < 0.001) and **A:8oxoG** lesion (estimate = −0.519 s.d., p = 0.006) repair activities were significantly reduced in patients who were assessed within one-month post-RT, but not in those evaluated later. In contrast, **LP-BER** (estimate = 0.474 s.d., p = 0.021) activity showed significant compensatory increase only in patients who returned for follow-up within 1-3 months post-RT, highlighting a dose- and time-dependent modulation of DNA repair pathway as following RT.

### 4.5. Effect modification of cancer stage and RT type on RT-induced DRC changes

NSCLC patients exhibited significant post-RT reductions in repair capacity for **U:G** lesion (estimate = −0.187 s.d., p = 0.034) and **A:8oxoG** lesion (estimate = −0.196 s.d., p = 0.038), whereas non-NSCLC patients showed no such changes [Figure 6A; Table S2]. A significant post-RT decrease in **A:8oxoG** lesion repair capacity was observed in patients with locally advanced cancer stage (estimate = −0.327 s.d., p = 0.008), but not in patients with earlier or metastatic stages [Figure 6B], potentially reflecting the higher RT doses and fractions regimens typically administered in this group. Additionally, a significant decline in **U:G** lesion repair capacity was detected exclusively in patients with widely metastatic disease (estimate = - 0.593 s.d., p = 0.043), aligning with the observation that individuals receiving palliative RT, who substantially overlapped with the metastatic group-also experienced a similar decline (estimate = −0.736 s.d., p = 0.058) [Figure 6C]. No significant modifiers were identified among patient demographics, including age, sex, or smoking history [Figure S5].

**Figure 6.**
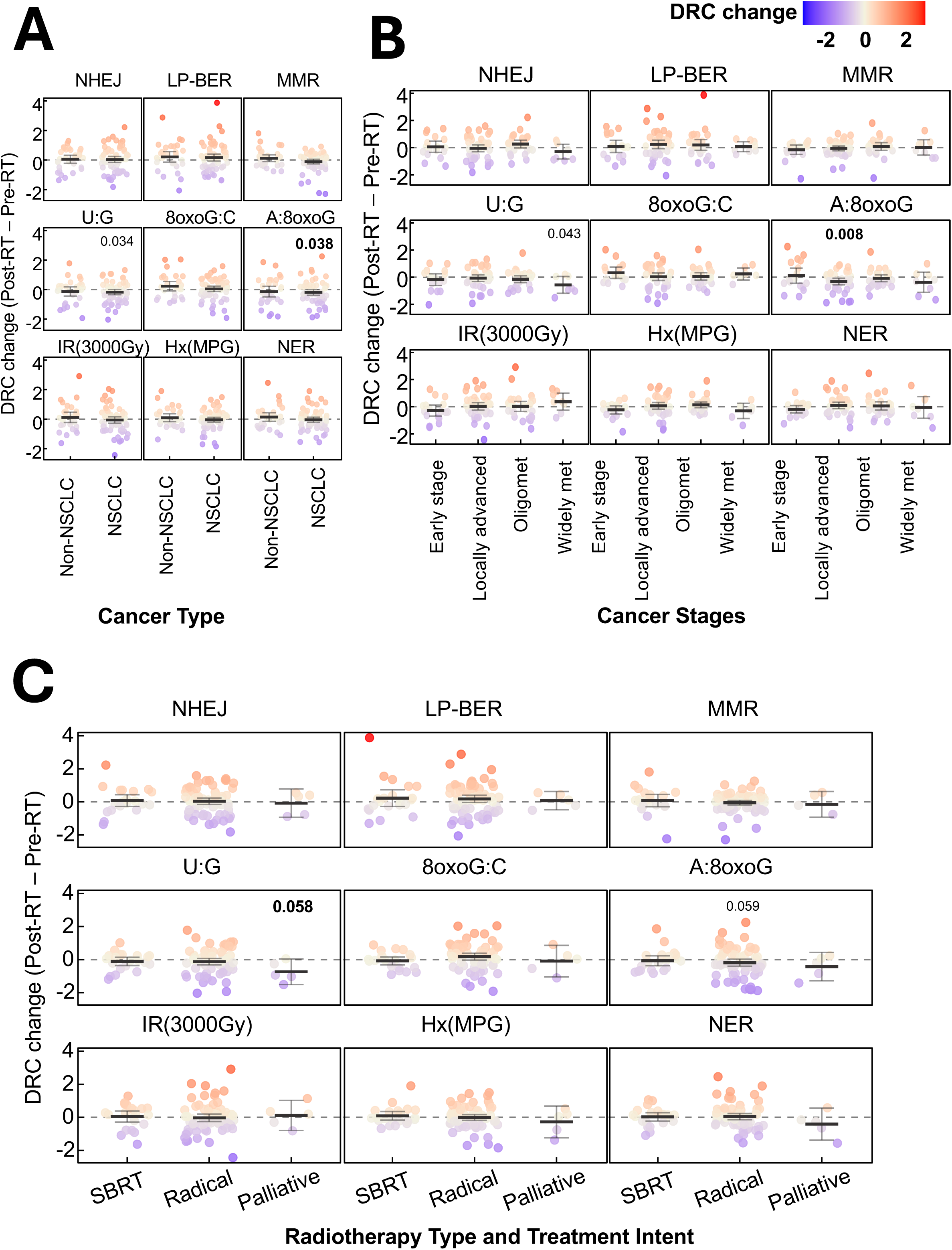
Effect modification of radiotherapy-induced DNA repair capacity changes by clinical characteristics. Radiotherapy (RT)-induced changes in DNA repair capacity (DRC), calculated as post-RT minus pre-RT values. Data were **(A)** stratified by cancer type: non-small cell lung cancer (NSCLC) and non-NSCLC, **(B)** stratified by cancer stage: early stage, locally advanced, oligometastatic, and widely metastatic, or **(C)** compared across radiotherapy types and treatment intent: stereotactic body radiation therapy (SBRT), radical, and palliative radiotherapy. Statistical significance (including borderline significance) of RT-induced DRC changes was assessed using linear mixed-effects models, with p-values indicated. Each subgroup is presented with mean values and 95% confidence intervals (error bars). The color gradient ranges from blue (indicating lower DRC changes) to red (indicating higher DRC changes).

Further stratification by RT type revealed that patients undergoing radical RT showed a borderline significant decrease in **A:8oxoG** lesion repair capacity (estimate = −0.203 s.d., p = 0.059), which reached statistically significant when restricted to the NSCLC subgroup (estimate = −0.243 s.d., p = 0.034). Similarly, a borderline significant decline in **U:G** lesion repair capacity was observed among NSCLC patients receiving radical RT (estimate = −0.190 s.d., p = 0.065). In contrast, non-NSCLC patients (post-RT) exhibited a significant increase was observed in repair capacity of **8oxoG:C** lesion (estimate = 0.506 s.d., p = 0.042) receiving radical RT. This collectively underscore the molecular impact of RT on DRC varies by cancer subtype and treatment intent with NSCLC patients showing heightened sensitivity in oxidative lesion repair pathways.

### 4.6. Altered DNA repair activities in lung cancer patients and their association with multiple primary tumors

We conducted a 1:1 matched case-control study, paring lung cancer patients with age-and sex-matched healthy controls, to assess differences in DRC. Compared to their matched healthy counterparts, lung cancer cases demonstrated a significantly elevated repair capacity for **NHEJ** (p < 0.001) and a reduced repair capacity for **A:8oxoG** lesions (p < 0.001) [Figure 7A]. These findings were corroborated by a subsequent covariate-adjusted group-wise analysis of all cases, which again demonstrated significant alterations in **NHEJ** (p < 0.001) and **A:8oxoG** lesions (p < 0.001) [Figure 7B, Table S5].

**Figure 7:**
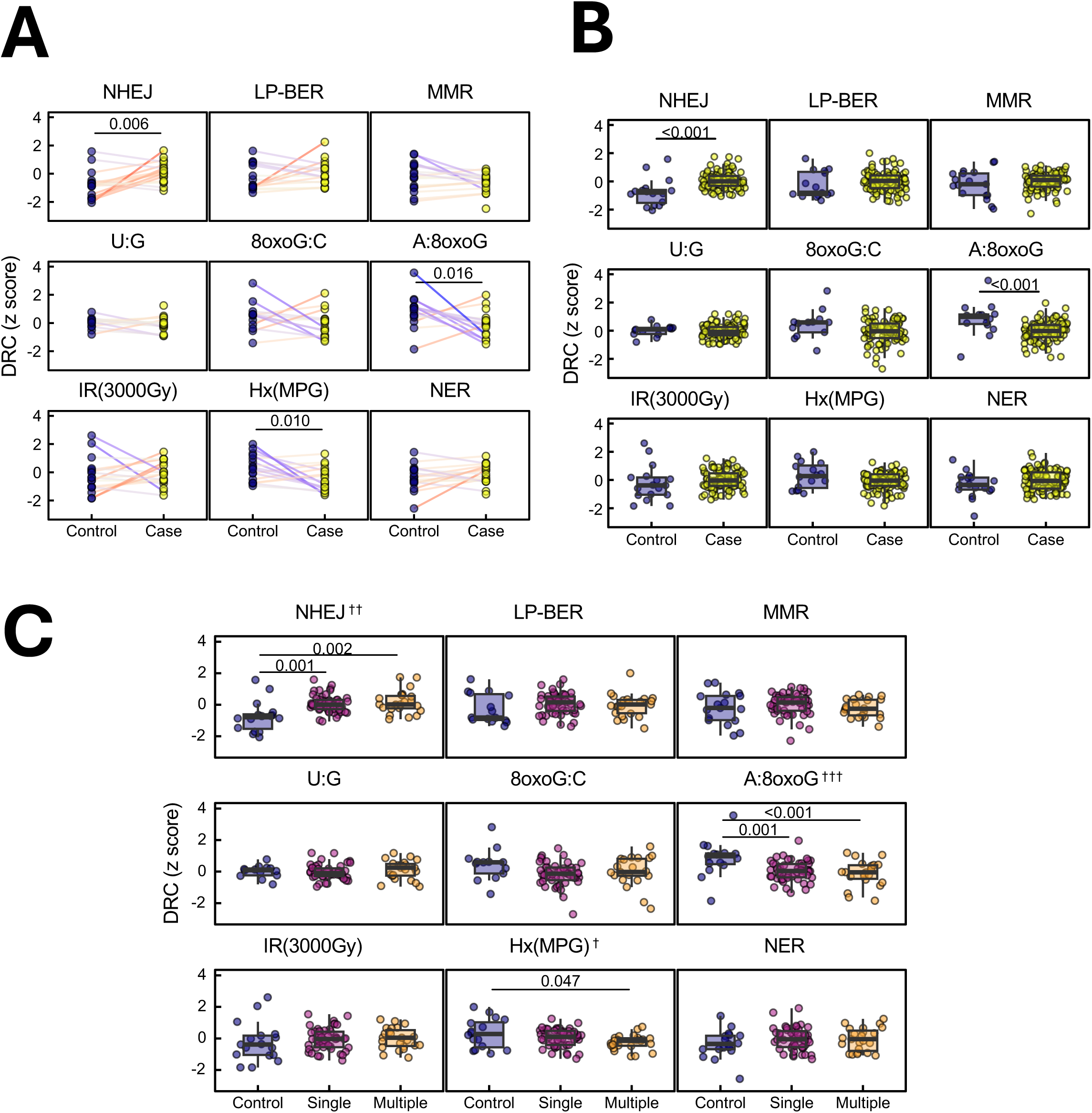
DNA repair capacity differences between lung cancer cases and healthy controls. **(A)**. Paired comparison of DRC between lung cancer patients (cases) and age- and sex-matched healthy controls, across multiple DNA repair pathways. Statistical significance was assessed using paired Student’s t-tests (n=18 matched pairs; case samples were post-therapy). **(B)** Comparison of DRC between the full set of lung cancer cases (n = 100) and healthy controls (n = 18). P-values were from generalized linear models adjusted for age and sex. (n = 18 control, n = 100 case, averaged for baseline and post-RT time points). **(C)** Stratification of DRC by the number of existing primary tumors: control (0), single (1), or multiple (2 or more). Statistical significance from generalized linear models is indicated by p-values. Trend tests were performed by modeling the number of primary tumors as a continuous variable to assess dose-response relationships with DRC. †p<0.05, ††p<0.01, †††p<0.001. (N=18 for control; N=52 for single, N=23 for multiple primaries).

We further investigated the potential dose-response relationship between the number of primary cancers and DRC, leveraging the diversity of our patient cohort and healthy controls. A significant positive trend was observed between the number of primary cancers and **NHEJ** repair capacity (trend test p = 0.006) and inversely associated with **A:8oxoG** lesion repair capacity (trend test p < 0.001) [Figure 7C, Table S5]. Moreover, cancer cases exhibited reduced **Hx:T** lesion repair capacity compared with their matched healthy controls [Figure 7A]. This reduction reached significance only in patients with multiple primary cancers (p = 0.047) and not in those with a single primary malignancy [p = 0.287], suggesting that increasing cancer severity might contribute to the observed declining trend (p = 0.041). Together, these findings highlight distinct lesion-specific alterations in DNA repair pathways in lung cancer patients, revealing that enhanced NHEJ but compromised oxidative lesion repair (A:8oxoG, Hx:T) characterize patients with greater cancer burden. Such patterns may reflect adaptative or selective repair reprogramming in tumorigenesis and underscore the potential of functional DRC profiling as a precision biomarker for cancer risk and progression monitoring.

## 5. Discussion

To date, no longitudinal, individual-based study has comprehensively assessed the full spectrum of DNA repair pathways and lesion-specific repair capacities due to technological limitations. This study, therefore, represents a novel and benchmark effort to profile DRC as a functional biomarker using advanced high throughout robustly validated technology in PBMCs ^10,13,25,30,37^, establishing a foundation for its integration into large scale clinical and population-based precision medicines studies. By evaluating multiple DNA repair pathways simultaneously in a diverse cohort of lung cancer patients, before and after RT, we have determined how DNA repair landscapes vary among individuals, depend on clinical variables, and change following RT [Figure 8]. When comparing patient characteristics with baseline DRC, we observed no significant associations with age, sex, or smoking status. These findings suggest that while demographics might influence DRC in healthy individuals or in case-control studies ^15,42–44^, they appear less predictive in this clinically heterogeneous lung cancer cohort. In our study, variation driven by cancer stage, prior treatment exposures, and comorbidities may outweigh variation that can be explained by demographics ^45–48^, which may only be evident in larger, more controlled studies. We also found no significant differences in baseline DRC between cancer types (NSCLC vs non-NSCLC), suggesting that intrinsic DRC might be driven by individual biological variability rather than influenced by cancer type. However, given the small and heterogeneous non-NSCLC group, our study may be underpowered to detect such differences, warranting cautious interpretation. Furthermore, our study included only 18 controls, limiting power for detecting differences.

**Figure 8.**
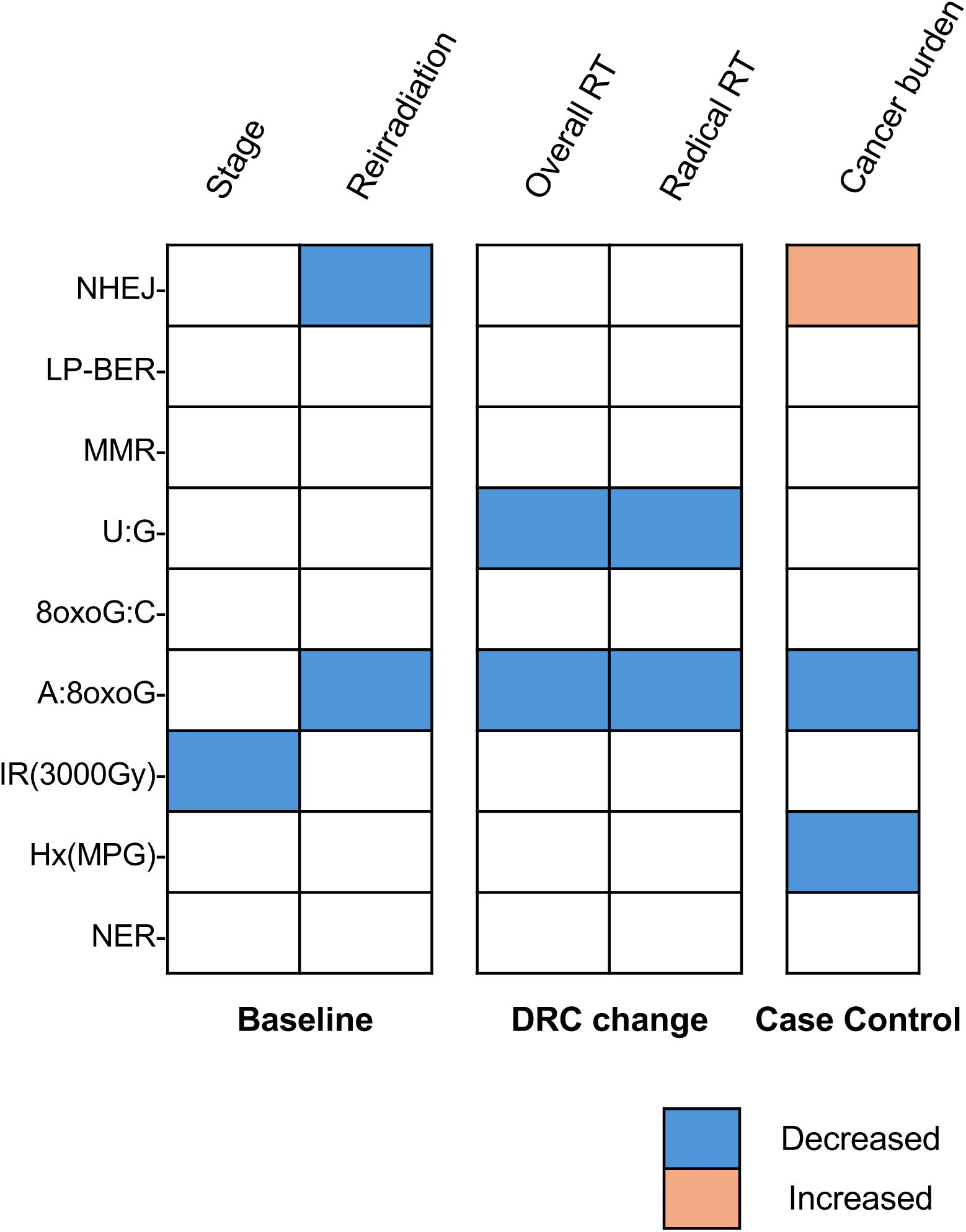
Association highlights between clinical factors and DNA repair capacity (DRC). We analyzed a range of patient characteristics association with each DNA repair assay measured at baseline and assessed their association with DRC changes following radiotherapy (RT). In addition, we examined the association between cancer burden (number of primary cancers) and DRC, using healthy controls as the reference group. Variables with significant associations are summarized in this heatmap. Blue indicates decreased DRC associated with higher levels of ordinal variables or “yes” in binary variables; orange indicates increased DRC associated with lower levels of ordinal variables.

We observed elevated baseline NHEJ repair capacity in cancer patients compared to healthy controls, suggesting a possible adaptive response to persistent DSB damage induced due to cancer development ^49–51^. Prior evidence indicated that although lower DSB repair capacity in healthy individuals may increase lung cancer risk, paradoxically higher DSB repair capacity during first-line treatments can be associated with more rapid progression ^50,52^. In our cohort, patients with widely metastatic stages and presenting with multiple primary cancers exhibited higher NHEJ than patients at early stage (Figure 3B) or with single primaries (Figure 7C), potentially reflecting heightened reliance on this repair pathway in the normal tissues of patients with advanced cancer. Although NHEJ may protect the genome in non-malignant tissues, its error-prone nature can lead to mutations that drive cancer progression and contribute to therapy resistance ^53^. Individuals with higher NHEJ repair capacity in their lymphocytes may also have higher NHEJ capacity in their tumor cells, which can lead to radio-resistance in lung cancer ^54–56^. Though DRC is frequently altered by somatic mutations and epigenetic changes in cancer, further work is needed to determine the extent to which DRC in cancer cells is associated with DRC in lymphocytes.

Notably, we observed a significant dose-dependent relationship between IR induced damage repair capacity and cancer stage, indicating that patients with more extensive disease might be more vulnerable to the health effects of IR-induced DNA damage. As we have discussed in a previous publication^37^, the dose needed to inactivate our relatively small (3.5kb) reporter plasmids are several orders of magnitude higher than the dose needed to kill a cell, but nevertheless induces the types and relative abundance of DNA lesions that occur at lower doses. This diminished DRC in advanced-stage patients may reflect a cumulative burden of systemic factors including chronic inflammation and impaired hematopoietic function ^57,58^. Since this effect was not evident for other pathway-specific measures, the mixture of DNA lesions or presence of clustered DNA lesions in the pMax_BFP_IR plasmid reporter may be more difficult to repair than isolated DNA lesions, thus making repair of this reporter plasmid more sensitive to minor changes in DRC.

Our results also showed strong correlations between NER and IR induced damage repair capacities both at baseline, post-RT visit, and in RT-induced changes, implying that either NER contributes significantly to the complex IR damage repair network ^59^, or NER is co-regulated with pathways that repair IR-induced DNA damage in quiescent lymphocytes. By contrast, IR induced damage repair capacity was not correlated with NHEJ, which remained largely stable following RT in our patient cohort, consistent with a previous report ^60^. While IR induces some DSBs ^20^, the vast majority of lesions induced by low-LET IR are base lesions, single strand breaks, and abasic sites ^61^. Consequently, IR is expected to require the action of several DNA repair pathways ^62^. NHEJ and LP-BER also showed weaker but significant positive correlations at both baseline and post-RT visits; notably, their RT-induced changes were also positively correlated, aligning with previous studies ^63^ and suggesting possible co-regulation of the two pathways. Additionally, patients with prior RT history showed consistently lower NHEJ than those without a similar history, though limited sample sizes preclude definitive conclusions. An overarching observation that applies to all pathways is that differences between individuals were generally much larger than the small shifts in DRC that occurred within individuals when comparing pre-versus post-RT. This points to the high reproducibility of FM-HCR assays in quiescent PBMCs, which permits the resolution of minor differences either within or between individuals ^13,37^.

Oxidative stress, arising from a variety of endogenous and exogenous sources, generates mutagenic oxidative lesions, including 7,8- dihydro-8-oxoguanine (8oxoG). Mis-incorporation of adenine opposite unrepaired 8oxoG lesions during replication can lead to CG->AT transversions ^64^. However the A:8oxoG mispair is recognized and repaired by the MutY glycosylase homologue (MUTYH) ^64^. MUTYH deficiency is associated with smoking status, higher increased mutation rates, and cancer risk ^65,66^. A central finding of our study is that A:8oxoG lesion repair capacity is a sensitive marker of RT response. Multiple RT variables, including treatment modality, dosage, fraction, follow-up intervals, and prior cancer history impacted A:8oxoG lesion repair dynamics. Specifically, we observed a significant reduction in A:8oxoG lesion repair capacity within one-month post-RT, followed by a return to baseline in later visits. This acute decrease may reflect overwhelming oxidative stress or transient suppression of key BER enzymes upon radiation exposure ^67,68^. The effect was more pronounced among patients receiving higher total radiation doses or more intensive fractionation, suggesting a dose-dependent link between RT-induced oxidative stress and oxidative lesion repair capacity ^69^. Further supporting a link between BER and physiological responses to treatment, we observed a reduction in U:G lesion repair that was also evident within one-month post-RT. The pronounced post-RT decline in A:8oxoG and U:G lesion repair, followed by a rebound to near-baseline levels, highlights the transient yet impactful nature of RT on BER pathways ^70–72^. In addition, a decrease in 8oxoG:C lesion repair capacity was evident in patients undergoing radical RT after restricting to non-NSCLC, highlighting heterogeneous effects of IR on the activity of different DNA glycosylases ^73^. Consistent with previous findings ^65,74,75^, we found persistent deficits in A:8oxoG lesion repair in lung cancer patients compared to controls, suggesting either pre-existing deficiency that may contribute to cancer development or cancer-driven alterations in the oxidative DNA damage responses, that could be influenced by tumor burden or metabolic changes. In addition, the dose-dependent decrease in repair of A:8oxoG lesions with the number of primary cancers further supported a strong link between cancer burden with oxidative damage response ^76^.

Notably, a low integrated DNA repair score, comprising key BER enzymes like OGG1, MPG and APE1 (OMA score), has previously been associated with lung cancer ^5^. Specifically, biochemical assays with cell-free extracts generated from PBMC indicated more efficient excision of Hx:T lesions and less efficient excision of 8oxoG:C lesion or incision of a tetrahydrofuran (THF) abasic site analog in lung cancer patients compared to cancer-free controls. In our study, FM-HCR indicated no significant differences between cases and controls in the repair of 8oxoG:C lesions or THF abasic site analog, and significantly less efficient repair of Hx:T among patients with multiple existing primary tumors when compared with cancer-free controls. Despite utilizing assays with the same DNA lesions, several significant differences between the studies may explain the differing results. First, the biochemical assays were run in extracts from mixed PBMCs that would reflect the average repair capacity of all cells present, whereas FM-HCR assays are dominated by signal from T cells. Second, whereas the biochemical assay is specific for THF incision by APE1, the fluorescent signal from the FM-HCR reporter GFP_THF is expected to reflect completion of LP-BER, since the single strand break intermediates downstream of abasic site incision are expected to block transcription. Third, FM-HCR reporters may be subject to the influence of transcription, which does not occur on the substrates used in biochemical assays. Fourth, the study population in Sevilya et al. comprised newly diagnosed lung cancer patients ^5^, while our patient cohort included individuals undergoing RT, many of whom had extensive prior treatment histories and more advanced stages. It is possible that the higher Hx:T lesion repair capacity observed in their study may decrease as cancer progresses, consistent with our finding of reduced Hx:T repair in patients with a greater burden of multiple primary tumors. However, the influence of cancer treatments is likely complex, as we observed varying baseline Hx:T lesion repair capacities among patients with different treatment histories, warranting further investigation in more controlled settings with longer follow-up. To mitigate potential confounding factors, these treatment variables were carefully adjusted for in our main analyses.

We furthermore noted that repair of A:8oxoG lesions, not measured in the previous studies, was significantly decreased in cancer patients. Reduced repair of the A:8oxoG lesion correlated with greater primary cancer burden, suggesting an additional risk for long-term genomic instability and secondary malignancies. A potential follow-up is to prospectively validate these observed relationships, as our current analysis identifies associations without definitive causal inference due to the heterogeneity of the case cohort and an inability to measure DRC prior to cancer development. While we did not directly evaluate cancer progression outcomes, these findings emphasize the importance of integrating both baseline and RT-induced DRC measurements into clinical evaluations ^35,77^.

We acknowledge several limitations that may influence the interpretation and generalizability of our findings. **First**, the use of cryopreserved PBMCs rather than fresh samples poses potential challenges ^33^. Although we adhered to standard FM-HCR protocols optimized for higher DRC reproducibility ^36^, cryopreservation may introduce variability in cellular function, sample feasibility, and batch effects. Our modified assay utilizing cryopreserved PBMCs has been published and demonstrated robustness, as cryopreservation did not significantly affect the DNA repair activity, cell viability, transfection efficiency and DRC rank order of individual. **Second**, due to sample limitations, our study lacks replicates for individual visits, limiting our ability to address random measurement errors within the dataset. However, repair capacity differences between individuals were highly reproducible at baseline and post-RT visits (Figure S6A-C) and robust to the significant but with smaller shifts in DRC that occurred during and after treatment. Reproducibility has furthermore been validated in previous work ^13^ and positive correlation trends between replicates in a random subset within this study (Figure S7A-B), and our association results were derived using regression models to mitigate random errors. Nevertheless, we cannot entirely exclude the potential bias toward the null introduced by these errors. Future studies should be designed in a manner that permits incorporating sample replicates to enhance data reliability and precision. **Third**, there is the potential selection bias due to the inclusion of patients based on sufficient isolated PBMCs cell count in cryovials, inherently excluding those with reduced cell counts who may exhibit altered responses to RT due to deficiencies in blood regeneration systems ^78,79^. Sensitivity analyses (Figure S7C) showed no significant correlation between cryopreserved PBMC counts and measured DRC, mitigating concerns that selection bias influenced the observed RT effects on DRC. However, the absence of external validation limits the broader applicability of these findings. The heterogeneity within the diverse non-NSCLC subgroup might compromise our ability to draw meaningful associations from themselves, and future research should prioritize the inclusion of independent cohorts in more controlled settings to confirm and extend these observations.

Our findings indicate that RT can induce systemic effects on DNA repair mechanisms, impacting both tumor cells and normal tissues, revealing both transient and persistent pathway-specific alterations in DRC. By conducting comprehensive longitudinal analyses within a clinically diverse patient cohort, we have characterized intrinsic stability and pronounced inter-individual variability in DRC, which is closely linked to critical clinical parameters such as cancer stage, radiation dose, and fractionation schedules. Particularly noteworthy is the elevated **NHEJ** in advanced cancer and RT-induced impairment of oxidative lesion repair (**A:8oxoG** lesion), highlighting the potential of DRC as a biomarker of treatment-related biological effects and radiation-induced toxicities that are driven, at least in part, by oxidative stress. These findings lay a robust foundation for translating functional DNA repair assays into clinical practice, potentially supporting personalized RT dosing and guiding the integration of DNA repair-modulating agents in combination therapies. Integration of high-throughput DRC phenotyping into the clinical workflow offers possible new strategies for refining risk stratification and informing the development of DNA repair modulating combination therapies and preventive measures for side effects. Future research will further elucidate the molecular mechanisms underlying observed changes in DRC, validate these functional biomarkers across broader clinical populations, and refine predictive models to advance personalized oncology and optimize patient outcomes.

## 7. Acknowledgement

We thank the clinical research coordinators Thomas Johnson, Sarah Caplan, Hejoo Kang, Seowon Kim, Maeve Dillon-Martin, Gulalai Shah, Teresia Perkins, John He, and Francois Georges, the patients, and the commercial sources (SMEBio, USA; Cellular Technology Limited, Ohio; Precision for Medicine, Massachusetts) that provided control PBMCs. We acknowledge Sujata Shah and her team at Dana Farber core facilities, Boston, MA, USA for their support for PBMC isolation and storage. The BROADBAND Project was possible in part by the generous donations of Stewart Clifford, Fredric Levin, and their families.

## 9. Author’s contributions

**SMT**: coordinated, designed, experimental work, data analysis & interpretation, and drafting the manuscript. **TZ**: experimental work data analysis & interpretation and drafting the manuscript**. MDM:** clinical data collection, sample collection and reviewed the manuscript. **PFD:** clinical data collection, regulatory management, sample collection and reviewed the manuscript. **CN:** sample collection and reviewed the manuscript. **DK:** supervision, conception and study design, sample collection, obtained funding and reviewed the manuscript. **ZDN:** supervision, conception and study design, obtained funding, reviewed the manuscript.

## 13. Data availability

The raw data generated and analyzed during this study are not publicly available due to patient privacy concerns. However, de-identified datasets can be obtained from the corresponding author upon reasonable request.

## 14. Code availability

R scripts used for statistical analyses and figures are deposited to GitHub: https://github.com/NagelLabHub/LC-FMHCR.

---

**Supplementary table 1.** Crude association between baseline DNA repair capacity and patient characteristics.

| Patient characteristics | NHEJ |  |  | LP-PBER |  |  | MMR |  |  | U:G |  |  |
| --- | --- | --- | --- | --- | --- | --- | --- | --- | --- | --- | --- | --- |
|  | beta | SE | p_value | beta | SE | p_value | beta | SE | p_value | beta | SE | p_value |
| Age (years) | -0.002 | 0.007 | 0.782 | 0.003 | 0.009 | 0.741 | -0.002 | 0.007 | 0.780 | -0.002 | 0.006 | 0.701 |
| Sex: femae vs male | 0.035 | 0.135 | 0.797 | 0.190 | 0.174 | 0.277 | 0.120 | 0.130 | 0.362 | -0.016 | 0.121 | 0.895 |
| Smoking: former vs never | 0.080 | 0.169 | 0.637 | -0.140 | 0.217 | 0.521 | 0.061 | 0.165 | 0.713 | 0.058 | 0.151 | 0.704 |
| Smoking: current vs never | 0.120 | 0.267 | 0.655 | -0.387 | 0.343 | 0.261 | -0.051 | 0.260 | 0.844 | 0.020 | 0.246 | 0.934 |
| Primary diagnosis: non-nsclc vs nsclc | 0.130 | 0.144 | 0.367 | -0.086 | 0.187 | 0.649 | -0.001 | 0.140 | 0.992 | 0.024 | 0.129 | 0.856 |
| Cancer stage: locally advanced vs early stage | **0.441** | **0.182** | **0.017** | 0.038 | 0.249 | 0.879 | -0.260 | 0.182 | 0.156 | 0.093 | 0.172 | 0.590 |
| Cancer stage: oligomet vs early stage | 0.036 | 0.196 | 0.854 | -0.024 | 0.268 | 0.928 | **-0.378** | **0.195** | **0.056** | -0.027 | 0.182 | 0.883 |
| Cancer stage: widely met vs early stage | 0.218 | 0.273 | 0.427 | 0.018 | 0.373 | 0.962 | -0.237 | 0.272 | 0.385 | 0.302 | 0.252 | 0.234 |
| Cancer stage: trend test | -0.024 | 0.079 | 0.764 | -0.010 | 0.103 | 0.922 | -0.111 | 0.076 | 0.147 | 0.036 | 0.071 | 0.613 |
| Number of primary diagnosis | 0.027 | 0.109 | 0.809 | -0.151 | 0.140 | 0.284 | -0.161 | 0.117 | 0.173 | 0.090 | 0.103 | 0.388 |
| Type: radical vs sbrt | 0.160 | 0.159 | 0.319 | -0.072 | 0.208 | 0.731 | 0.063 | 0.153 | 0.680 | 0.099 | 0.142 | 0.489 |
| Type: palliative vs sbrt | -0.039 | 0.327 | 0.905 | 0.157 | 0.426 | 0.714 | **0.582** | **0.312** | **0.066** | 0.205 | 0.288 | 0.479 |
| Prior radiotherapy (any) | **-0.367** | **0.182** | **0.047** | -0.356 | 0.241 | 0.143 | -0.094 | 0.181 | 0.604 | -0.074 | 0.165 | 0.656 |
| Prior radiotherapy (within 6 months) | **-0.533** | **0.274** | **0.055** | -0.569 | 0.360 | 0.118 | 0.161 | 0.271 | 0.553 | -0.205 | 0.246 | 0.406 |
| Prior chemotherapy | 0.109 | 0.145 | 0.455 | 0.117 | 0.189 | 0.538 | 0.113 | 0.141 | 0.426 | 0.174 | 0.129 | 0.181 |
| Prior immunotherapy | -0.102 | 0.197 | 0.606 | 0.150 | 0.256 | 0.559 | -0.071 | 0.191 | 0.711 | -0.153 | 0.174 | 0.381 |
| Prior targeted therapy | -0.114 | 0.181 | 0.531 | 0.200 | 0.235 | 0.396 | -0.204 | 0.174 | 0.245 | 0.150 | 0.159 | 0.349 |
| Race: black vs white | -0.328 | 0.470 | 0.487 | 0.167 | 0.619 | 0.788 | **0.972** | **0.455** | **0.035** | 0.323 | 0.422 | 0.445 |
| Race: asian vs white | -0.334 | 0.386 | 0.388 | 0.044 | 0.508 | 0.930 | -0.125 | 0.373 | 0.739 | 0.145 | 0.346 | 0.676 |

| Patient characteristics | 8oxoG:C |  |  | A:8oxoG |  |  | IR(3000Gy) |  |  | Hx(MPG) |  |  | NER |  |  |
| --- | --- | --- | --- | --- | --- | --- | --- | --- | --- | --- | --- | --- | --- | --- | --- |
|  | beta | SE | p_value | beta | SE | p_value | beta | SE | p_value | beta | SE | p_value | beta | SE | p_value |
| Age (years) | -0.015 | 0.009 | 0.108 | 0.001 | 0.009 | 0.899 | 0.000 | 0.009 | 0.977 | -0.010 | 0.007 | 0.198 | 0.000 | 0.009 | 0.974 |
| Sex: femae vs male | -0.131 | 0.186 | 0.483 | 0.208 | 0.175 | 0.236 | 0.141 | 0.169 | 0.404 | -0.203 | 0.143 | 0.159 | 0.233 | 0.168 | 0.170 |
| Smoking: former vs never | 0.112 | 0.230 | 0.626 | 0.211 | 0.216 | 0.331 | 0.038 | 0.213 | 0.857 | 0.093 | 0.182 | 0.609 | 0.155 | 0.215 | 0.471 |
| Smoking: current vs never | -0.394 | 0.394 | 0.320 | 0.320 | 0.340 | 0.350 | -0.113 | 0.349 | 0.747 | -0.015 | 0.287 | 0.959 | 0.244 | 0.338 | 0.472 |
| Primary diagnosis: non-nsclc vs nsclc | 0.262 | 0.199 | 0.192 | -0.076 | 0.188 | 0.687 | 0.043 | 0.182 | 0.813 | -0.059 | 0.155 | 0.702 | -0.057 | 0.182 | 0.756 |
| Cancer stage: locally advanced vs early stage | 0.146 | 0.269 | 0.590 | 0.368 | 0.245 | 0.137 | **-0.490** | **0.232** | **0.037** | -0.285 | 0.202 | 0.161 | -0.312 | 0.237 | 0.192 |
| Cancer stage: oligomet vs early stage | -0.179 | 0.287 | 0.534 | 0.099 | 0.263 | 0.708 | -0.409 | 0.249 | 0.104 | -0.118 | 0.217 | 0.589 | -0.419 | 0.256 | 0.105 |
| Cancer stage: widely met vs early stage | 0.045 | 0.410 | 0.913 | 0.205 | 0.367 | 0.577 | **-0.796** | **0.347** | **0.024** | 0.151 | 0.303 | 0.620 | -0.484 | 0.356 | 0.178 |
| Cancer stage: trend test | -0.076 | 0.113 | 0.501 | 0.005 | 0.103 | 0.960 | **-0.191** | **0.097** | **0.053** | 0.046 | 0.085 | 0.592 | -0.163 | 0.099 | 0.103 |
| Number of primary diagnosis | 0.107 | 0.162 | 0.512 | -0.185 | 0.132 | 0.167 | 0.047 | 0.133 | 0.725 | **-0.201** | **0.114** | **0.081** | -0.032 | 0.136 | 0.812 |
| Type: radical vs sbrt | 0.134 | 0.219 | 0.543 | 0.273 | 0.207 | 0.190 | 0.036 | 0.201 | 0.859 | -0.170 | 0.169 | 0.318 | -0.002 | 0.202 | 0.993 |
| Type: palliative vs sbrt | 0.274 | 0.443 | 0.538 | 0.312 | 0.423 | 0.462 | -0.078 | 0.411 | 0.850 | 0.376 | 0.347 | 0.282 | 0.360 | 0.413 | 0.386 |
| Prior radiotherapy (any) | 0.192 | 0.254 | 0.451 | -0.304 | 0.241 | 0.210 | -0.208 | 0.233 | 0.376 | 0.209 | 0.200 | 0.300 | -0.051 | 0.235 | 0.827 |
| Prior radiotherapy (within 6 months) | -0.204 | 0.380 | 0.592 | **-0.664** | **0.358** | **0.067** | 0.130 | 0.351 | 0.711 | 0.302 | 0.301 | 0.318 | 0.191 | 0.351 | 0.587 |
| Prior chemotherapy | 0.196 | 0.202 | 0.336 | 0.158 | 0.189 | 0.405 | -0.206 | 0.183 | 0.263 | **0.312** | **0.153** | **0.044** | -0.013 | 0.184 | 0.943 |
| Prior immunotherapy | -0.200 | 0.268 | 0.458 | -0.073 | 0.257 | 0.776 | 0.043 | 0.247 | 0.863 | **0.483** | **0.207** | **0.022** | 0.103 | 0.249 | 0.679 |
| Prior targeted therapy | -0.028 | 0.253 | 0.913 | 0.286 | 0.234 | 0.225 | 0.087 | 0.227 | 0.701 | -0.207 | 0.194 | 0.288 | -0.158 | 0.228 | 0.491 |
| Race: black vs white | -0.611 | 0.640 | 0.342 | -0.215 | 0.629 | 0.734 | 0.740 | 0.584 | 0.208 | **0.842** | **0.503** | **0.098** | 0.669 | 0.598 | 0.266 |
| Race: asian vs white | -0.471 | 0.526 | 0.373 | 0.305 | 0.517 | 0.556 | 0.300 | 0.480 | 0.534 | -0.112 | 0.413 | 0.787 | -0.477 | 0.491 | 0.333 |

Statistics are obtained from univariate linear regression models.
Abbreviations: SE = standard error.

**Supplementary table 2.**
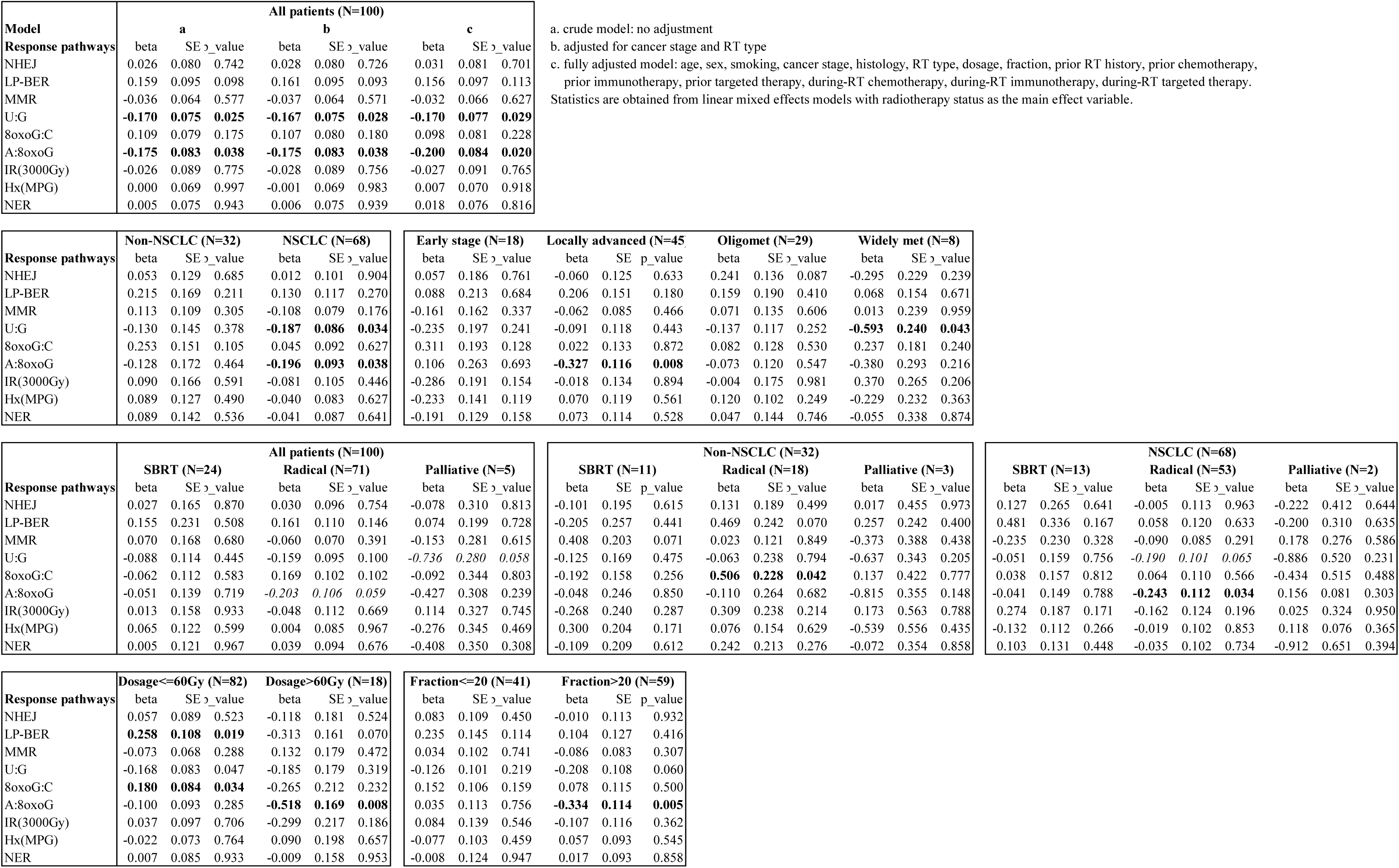
Radiotherapy response in DNA repair capacity and by patient status.

**Supplementary table 3.**
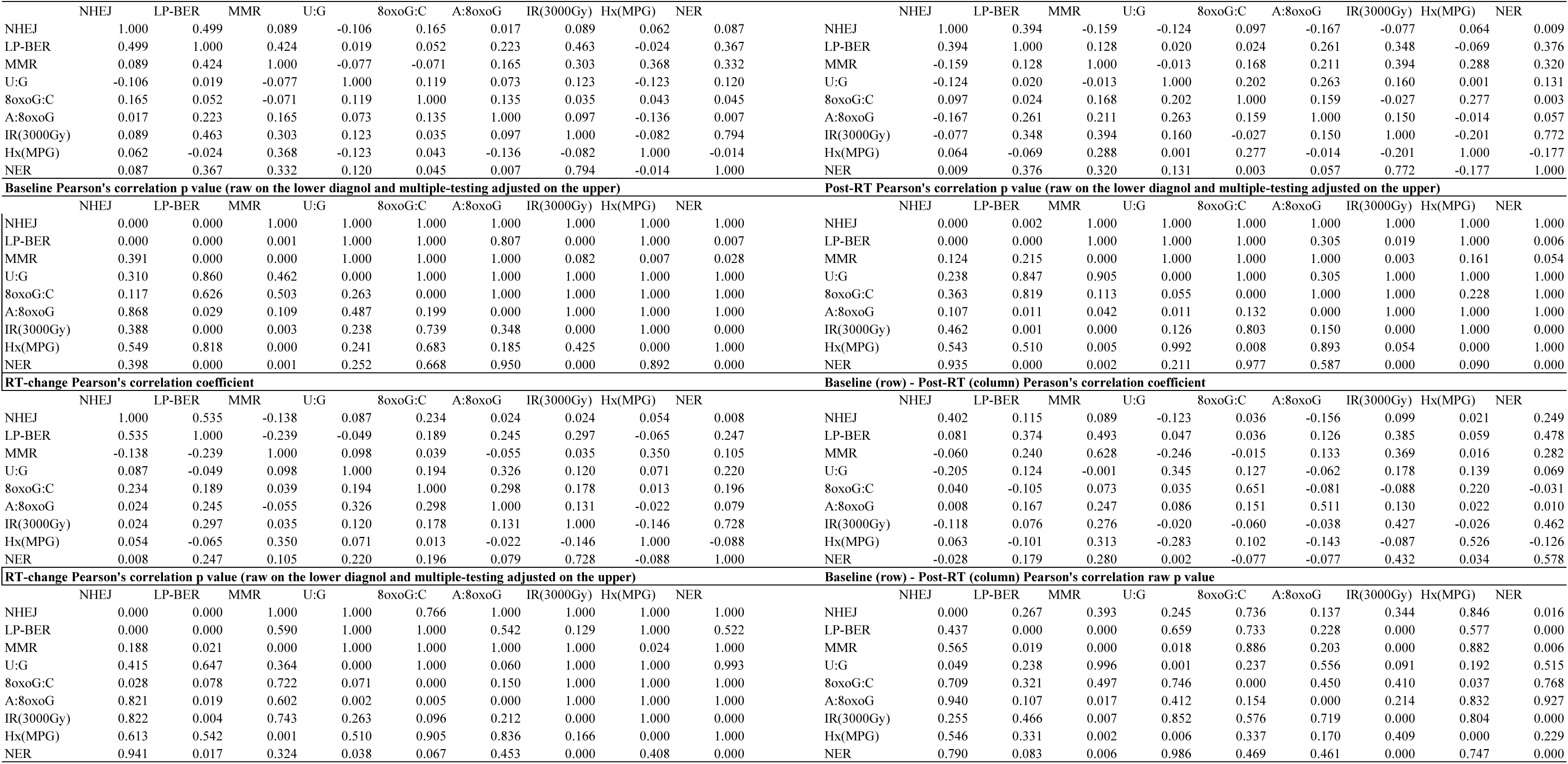
Correlations of DNA repair capacity measurements at baseline and at post-radiotherapy visit.

**Supplementary table 4.** Radiotherapy response in DNA repair capacity by follow-up months.

| Post-RT month groups | NHEJ |  |  | LP-PBER |  |  | MMR |  |  | U:G |  |  |
| --- | --- | --- | --- | --- | --- | --- | --- | --- | --- | --- | --- | --- |
|  | beta | SE | p_value | beta | SE | p_value | beta | SE | p_value | beta | SE | p_value |
| Within 1 month (N=18) | 0.172 | 0.166 | 0.300 | **0.474** | **0.202** | **0.021** | -0.025 | 0.146 | 0.864 | **-0.527** | **0.153** | **0.001** |
| 1-3 months (N=35) | 0.146 | 0.115 | 0.207 | 0.176 | 0.142 | 0.218 | -0.071 | 0.100 | 0.479 | -0.171 | 0.104 | 0.102 |
| 3-6 months (N=25) | -0.055 | 0.134 | 0.684 | 0.099 | 0.165 | 0.550 | 0.011 | 0.117 | 0.926 | -0.006 | 0.126 | 0.960 |
| 6-12 months (N=14) | -0.026 | 0.175 | 0.884 | -0.041 | 0.214 | 0.850 | 0.013 | 0.153 | 0.931 | -0.162 | 0.157 | 0.304 |
| >12 months (N=5) | **-0.705** | **0.288** | **0.015** | -0.101 | 0.351 | 0.774 | -0.197 | 0.253 | 0.438 | 0.145 | 0.257 | 0.573 |

| Post-RT month groups | 8oxoG:C |  |  | A:8oxoG |  |  | IR(3000Gy) |  |  | Hx(MPG) |  |  | NER |  |  |
| --- | --- | --- | --- | --- | --- | --- | --- | --- | --- | --- | --- | --- | --- | --- | --- |
|  | beta | SE | p_value | beta | SE | p_value | beta | SE | p_value | beta | SE | p_value | beta | SE | p_value |
| Within 1 month (N=18) | 0.222 | 0.189 | 0.242 | **-0.519** | **0.186** | **0.006** | 0.094 | 0.197 | 0.635 | -0.118 | 0.155 | 0.448 | 0.292 | 0.171 | 0.089 |
| 1-3 months (N=35) | 0.151 | 0.120 | 0.212 | -0.117 | 0.124 | 0.348 | -0.084 | 0.133 | 0.530 | 0.058 | 0.103 | 0.576 | -0.032 | 0.113 | 0.780 |
| 3-6 months (N=25) | 0.109 | 0.149 | 0.469 | -0.076 | 0.149 | 0.611 | 0.043 | 0.158 | 0.785 | -0.155 | 0.126 | 0.220 | 0.007 | 0.136 | 0.957 |
| 6-12 months (N=14) | 0.066 | 0.192 | 0.734 | -0.161 | 0.190 | 0.398 | -0.090 | 0.202 | 0.655 | 0.162 | 0.158 | 0.307 | -0.112 | 0.174 | 0.521 |
| >12 months (N=5) | -0.375 | 0.307 | 0.224 | -0.102 | 0.314 | 0.746 | -0.118 | 0.332 | 0.723 | 0.177 | 0.261 | 0.498 | -0.242 | 0.288 | 0.403 |

Statistics are obtained from linear mixed effect models with the post-RT month groups as the main effect variables.

**Supplementary table 5.** Case control comparison for DNA repair capacity.

| **DRC** | **Case (N=18) vs Control (N=18)** |  |  | **Case (N=100) vs Control (N=18)\*** |  |  |
| --- | --- | --- | --- | --- | --- | --- |
|  | beta | SE | p_value | beta | SE | p_value |
| NHEJ | **0.850** | **0.277** | **0.006** | **0.810** | **0.195** | **<0.001** |
| LP-BER | 0.388 | 0.289 | 0.196 | 0.302 | 0.228 | 0.188 |
| MMR | -0.388 | 0.224 | 0.103 | 0.147 | 0.198 | 0.458 |
| U:G | -0.094 | 0.166 | 0.581 | -0.050 | 0.152 | 0.742 |
| 8oxoG:C | -0.535 | 0.338 | 0.125 | -0.390 | 0.258 | 0.134 |
| A:8oxoG | **-0.885** | **0.347** | **0.016** | **-1.162** | **0.220** | **<0.001** |
| IR(3000Gy) | 0.135 | 0.341 | 0.694 | 0.088 | 0.217 | 0.686 |
| Hx(MPG) | **-0.798** | **0.291** | **0.010** | -0.238 | 0.186 | 0.205 |
| NER | 0.259 | 0.232 | 0.281 | 0.157 | 0.198 | 0.428 |

| **DRC** | **Trend test by number of primary** |  |  | **Single primary vs control** |  |  | **Multi primaries vs control** |  |  |
| --- | --- | --- | --- | --- | --- | --- | --- | --- | --- |
|  | beta | SE | p_value | beta | SE | p_value | beta | SE | p_value |
| NHEJ | **0.367** | **0.130** | **0.006** | **0.773** | **0.218** | **0.001** | **0.837** | **0.257** | **0.002** |
| LP-BER | 0.027 | 0.144 | 0.851 | 0.328 | 0.246 | 0.186 | 0.132 | 0.291 | 0.653 |
| MMR | -0.051 | 0.136 | 0.709 | 0.183 | 0.227 | 0.424 | -0.05 | 0.273 | 0.855 |
| U:G | 0.067 | 0.093 | 0.470 | -0.038 | 0.162 | 0.813 | 0.104 | 0.19 | 0.584 |
| 8oxoG:C | -0.092 | 0.162 | 0.571 | -0.516 | 0.276 | 0.065 | -0.298 | 0.325 | 0.362 |
| A:8oxoG | **-0.532** | **0.141** | **<0.001** | -1.072 | 0.232 | **<0.001** | -1.205 | 0.274 | **<0.001** |
| IR(3000Gy) | 0.078 | 0.144 | 0.588 | 0.087 | 0.246 | 0.725 | 0.159 | 0.293 | 0.59 |
| Hx(MPG) | **-0.233** | **0.112** | **0.041** | -0.205 | 0.191 | 0.287 | **-0.461** | **0.229** | **0.047** |
| NER | 0.079 | 0.127 | 0.537 | 0.184 | 0.216 | 0.397 | 0.181 | 0.258 | 0.486 |

\* adjusted for age and sex
Statistics are obtained from linear regression models.

**Supplementary figure 1:**
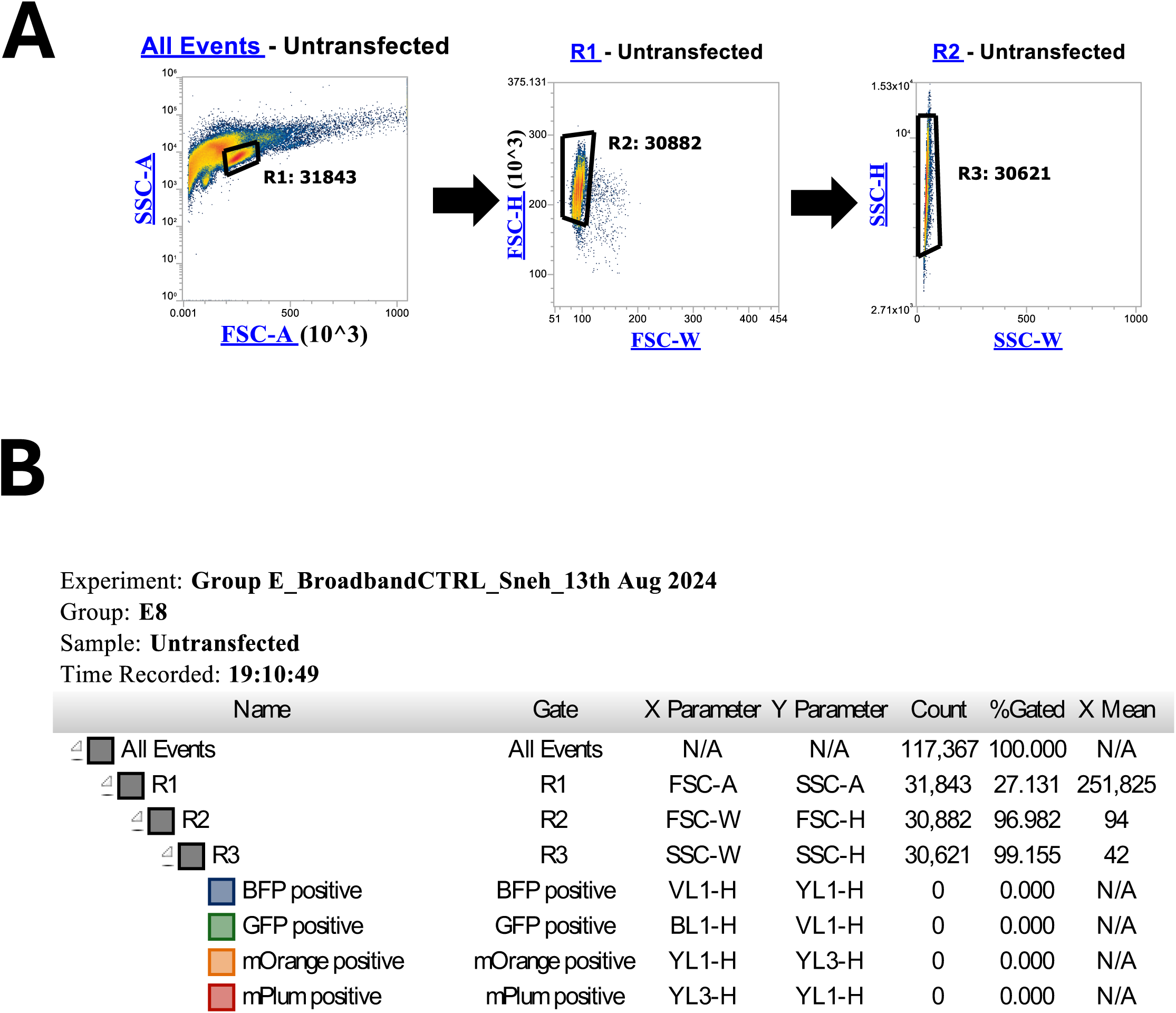
Hierarchy of cell selection in flow cytometry. **(A)** Scatter plots illustrating gates used for identifying singlets using flow cytometry in forward scatter (FSC) versus side scatter (SSC). **(B)** Gating hierarchy.

**Supplementary figure 2:**
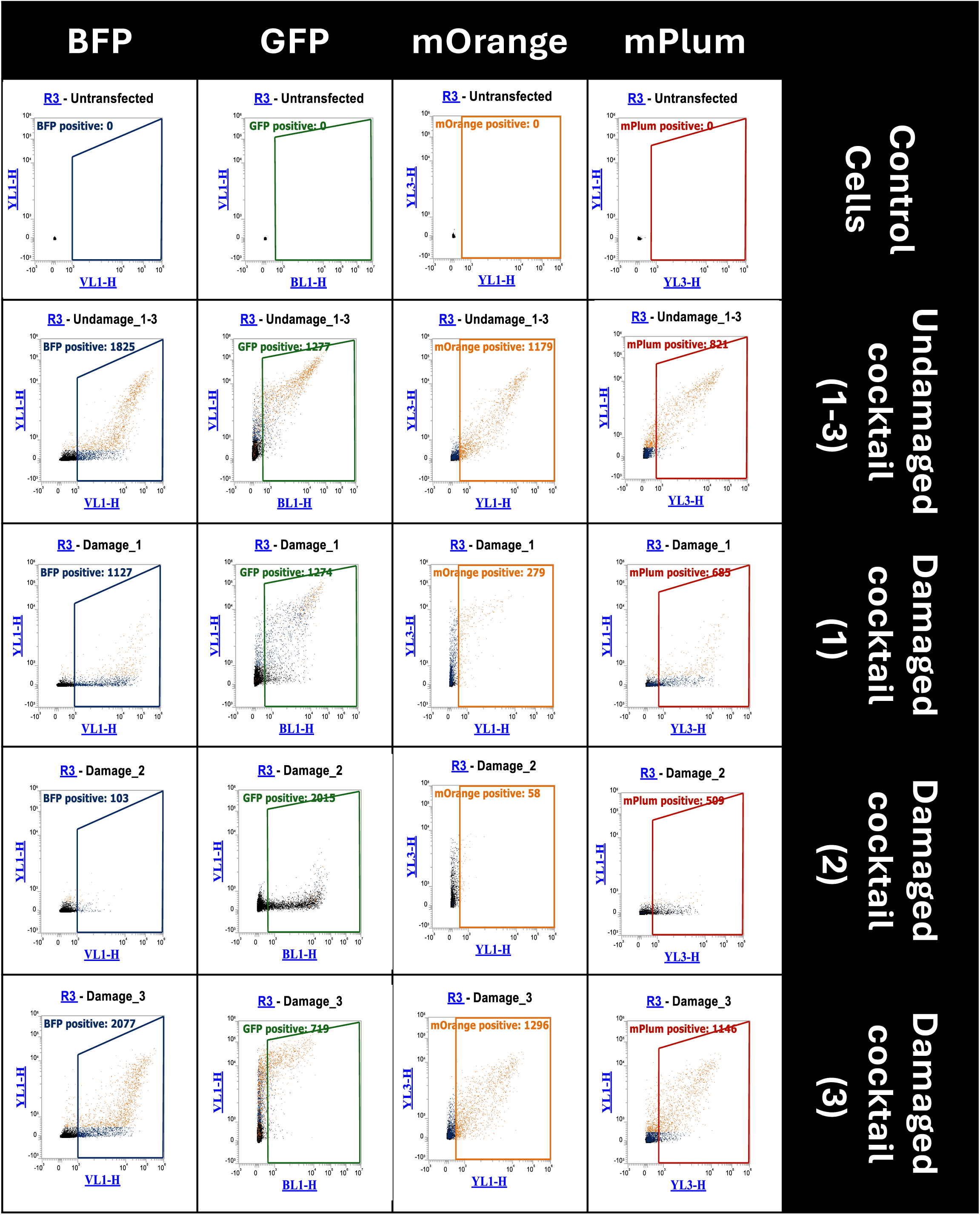
Representative flow cytometry scatter plots for PBMCs transfected with the reporter plasmid cocktails described in Table 1.

**Supplementary figure 3:**
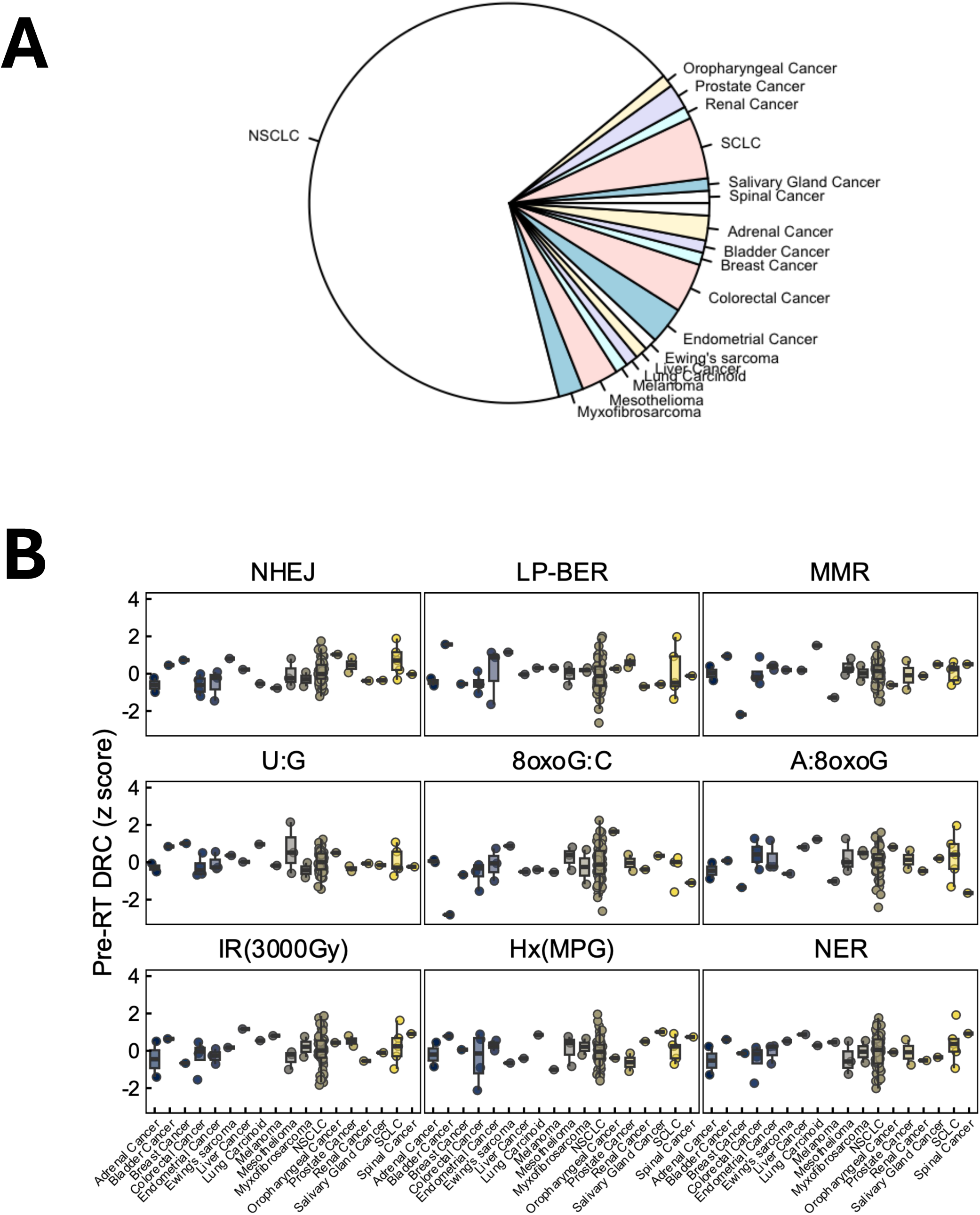
Cancer type distribution (A) and associated baseline DNA repair capacity variations (B).

**Supplementary figure 4:**
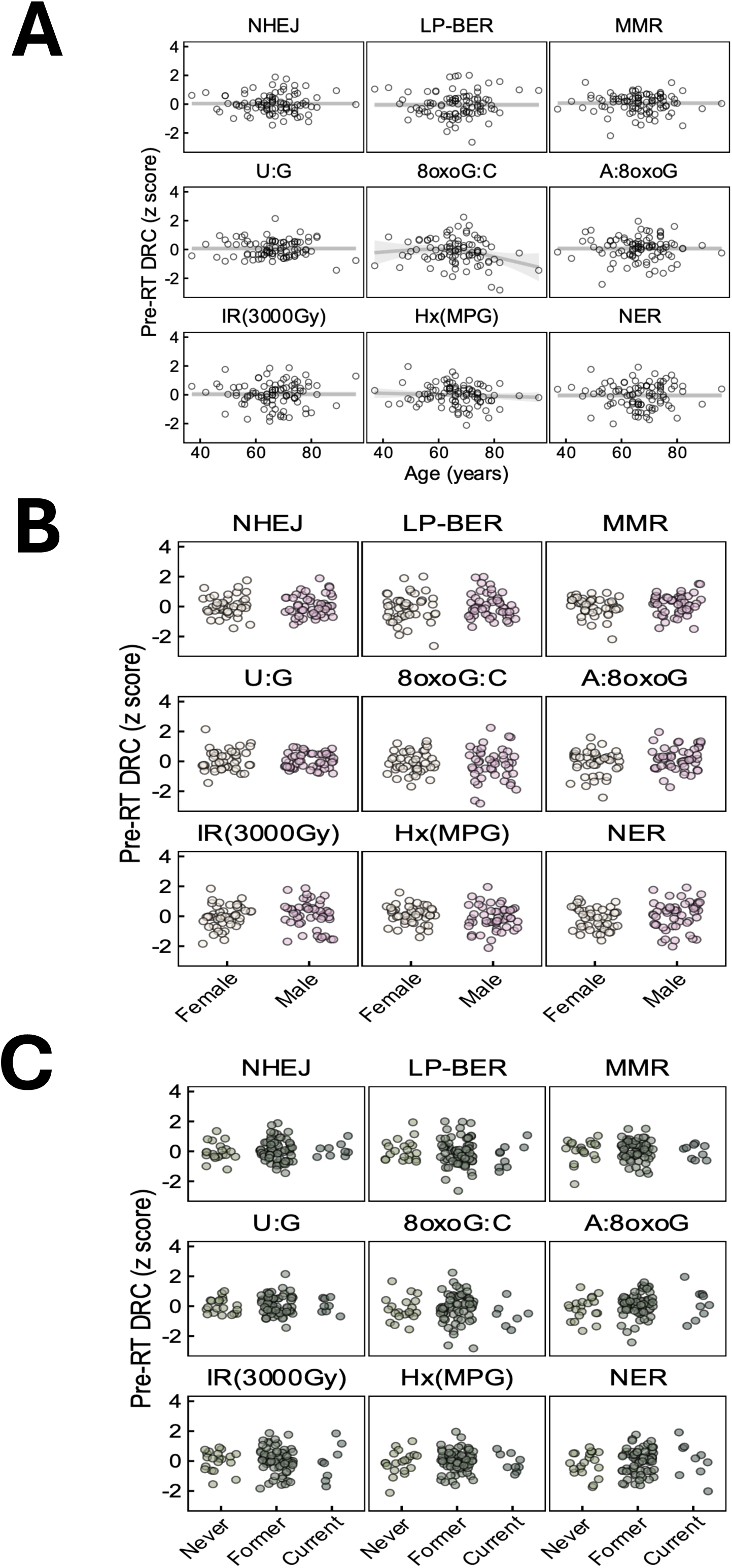
Baseline DNA repair capacity by demographics. Z-scored DNA repair capacity (DRC) at the pre-radiotherapy (baseline) timepoint is evaluated in relation to patient demographics: **(A)** age (in years), **(B)** sex (male, female) and **(C)** smoking status (never, former, current).

**Supplementary figure 5:**
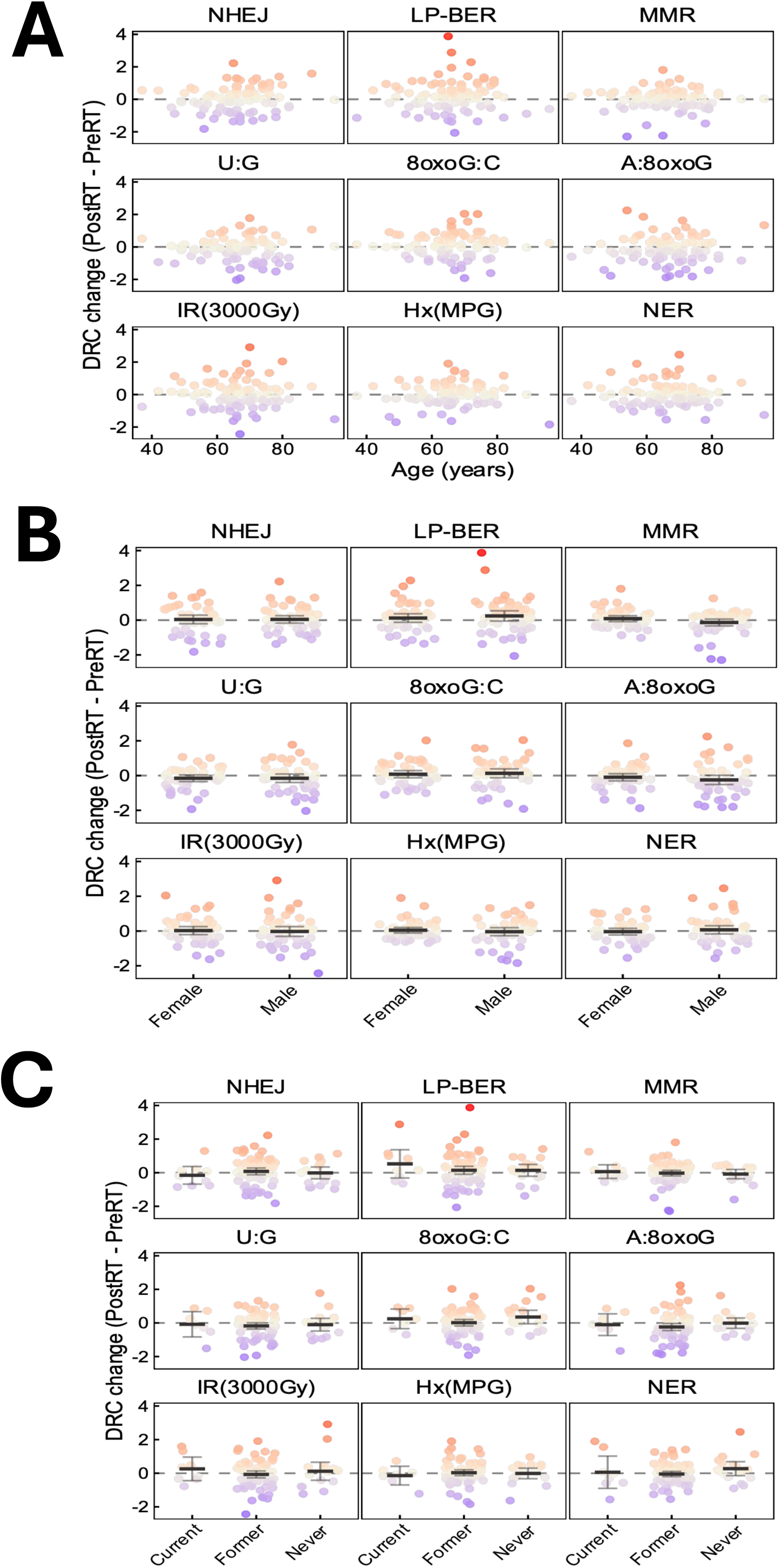
Radiotherapy-induced changes in DNA repair capacity by demographics. Changes in DRC (post-RT minus baseline, z-scored) are assessed in relation to: **(A)** age (in years), **(B)** sex (male, female) and **(C)** smoking status (never, former, current).

**Supplementary figure 6:**
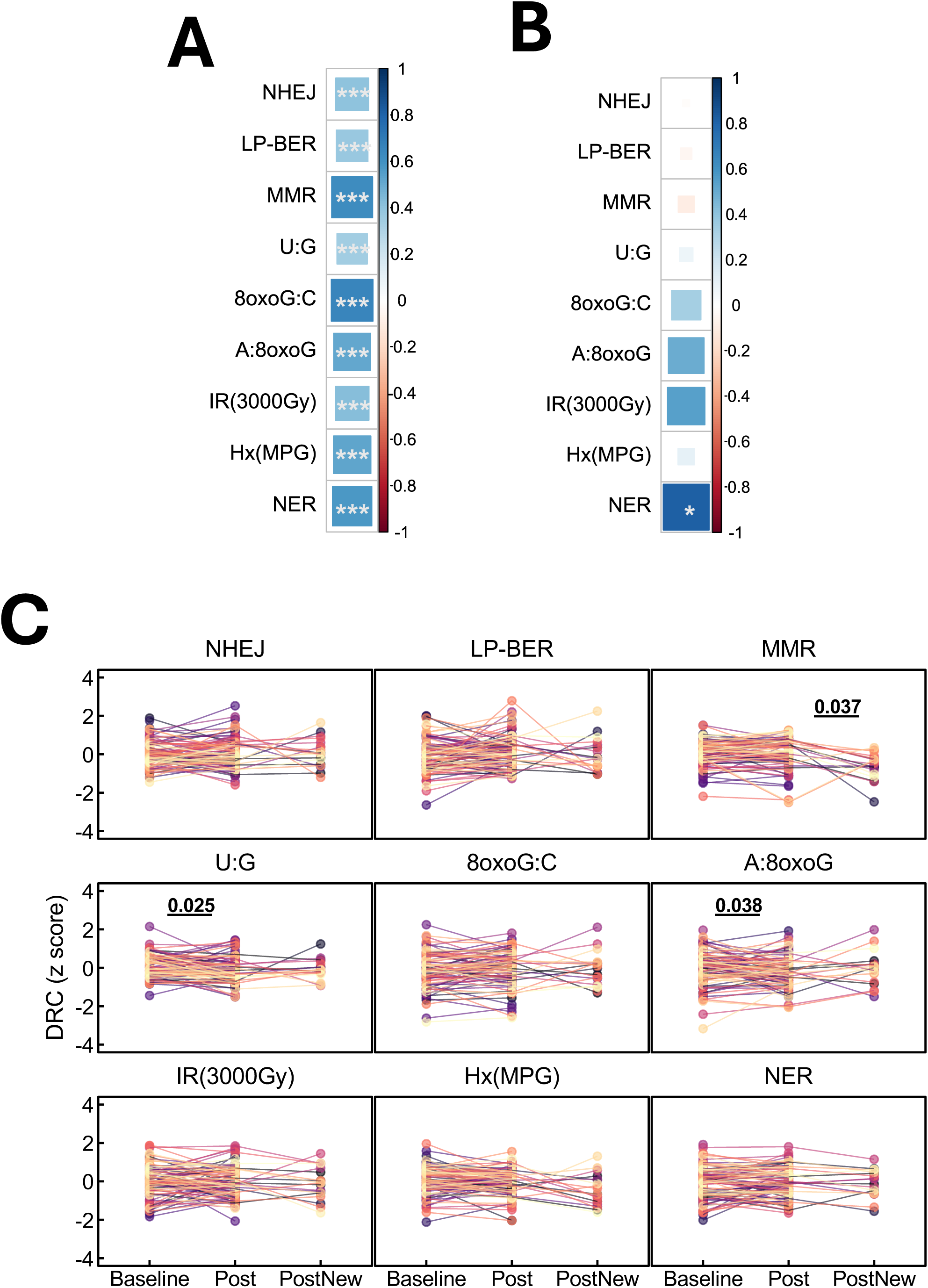
Correlation and paired comparisons of DNA repair capacity across timepoints in patient samples. **(A)** Correlations between pre-RT and post-RT samples within the same patients. **(B)** Correlations between two independent post-RT samples collected from the same patients at distinct follow-up timepoints. **(C)** Paired comparisons of DNA repair capacity across pre-RT, post-RT, and additional post-RT visits (PostNew) within the same patients. Statistical significance of pairwise comparisons is indicated by p-values derived from linear mixed-effects models.

**Supplementary figure 7.**
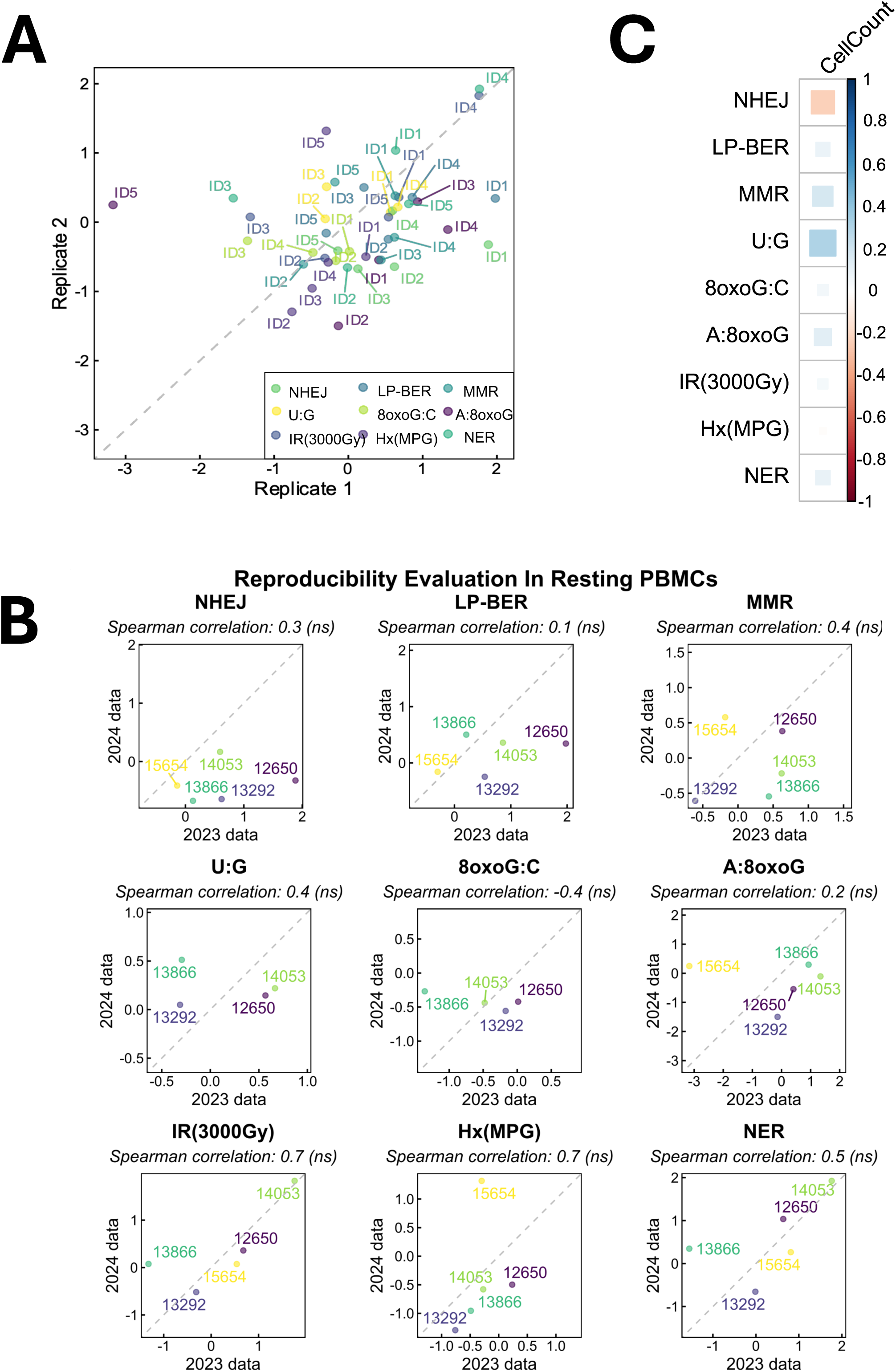
Technical reproducibility and assay validation. **(A)** Reproducibility of DRC assays assessed in a subset of patients (IDs 1-5) with pre-RT (baseline) samples measured in two separate years: DRC measurements (z-scores) at 2023 (Replicate 1) and 2024 (Replicate 2). Each point represents one measurement per replicate, colored by assay type. **(B)** Per-assay reproducibility across NHEJ, LP-BER, MMR, U:G, 8oxoG:C, A:8oxoG, IR(3000Gy), Hx(MPG), and NER, was assessed in replicate samples from case individuals. Replicates were processed in two different years: 2023 (Replicate 1) and 2024 (Replicate 2). Each panel shows z-scored DRC values for the two replicates, along with Spearman correlation coefficients and significance (ns = not significant). **(C)** Spearman correlation between DRC and PBMC cell count across collected samples.

